# Leveraging large-scale multi-omics to identify therapeutic targets from genome-wide association studies

**DOI:** 10.1101/2023.11.01.23297926

**Authors:** Samuel Lessard, Michael Chao, Kadri Reis, FinnGen, Estonian Biobank Research Team, Mathieu Beauvais, Deepak K. Rajpal, Srinivas Shankara, Jennifer Sloane, Priit Palta, Katherine Klinger, Emanuele de Rinaldis, Shameer Khader, Clément Chatelain

**Affiliations:** Precision Medicine & Computational Biology, Sanofi US, Cambridge, MA, USA; Estonian Genome Centre, Institute of Genomics, University of Tartu, Tartu, Estonia; Digital R&D Data & Computational Sciences, Sanofi, Chilly-Mazarin, France; Translational Sciences, Sanofi US, Framingham, MA, USA; Pre-Clinical and Translational Sciences, Takeda, MA, USA; Immunology & Inflammation Development, Sanofi US, Cambridge, MA, USA; Genetics Research, Sanofi US, Cambridge, MA, USA

**Author notes:** Employee of Sanofi US at the time of study. These authors contributed equally (Shameer Khader, Clément Chatelain). Corresponding author: Samuel Lessard.

**Keywords:** Genome-wide association study, molecular quantitative trait loci, causal inference, therapeutic targets, interleukin 6, polymyalgia rheumatica, mendelian randomization

## Abstract

**BACKGROUND**: Therapeutic targets supported by genetic evidence from genome-wide association studies (GWAS) show higher probability of success in clinical trials. GWAS is a powerful approach to identify links between genetic variants and phenotypic variation; however, identifying the genes driving associations identified in GWAS remains challenging. Integration of molecular quantitative trait loci (molQTL) such as expression QTL (eQTL) using mendelian randomization (MR) and colocalization analyses can help with the identification of causal genes. Careful interpretation remains warranted because eQTL can affect the expression of multiple genes within the same locus. **METHODS**: We used a combination of genomic features that include variant annotation, activity-by-contact maps, MR, and colocalization with molQTL to prioritize causal genes across 4,611 disease GWAS and meta-analyses from biobank studies, namely FinnGen, Estonian Biobank and UK Biobank. **RESULTS**: Genes identified using this approach are enriched for gold standard causal genes and capture known biological links between disease genetics and biology. In addition, we find that eQTLs colocalizing with GWAS are statistically enriched for corresponding disease-relevant tissues. We show that predicted directionality from MR is generally consistent with matched drug mechanism of actions (>78% for approved drugs). Compared to the nearest gene mapping method our approach also shows a higher enrichment in approved therapeutic targets (risk ratio 1.38 vs 2.06). Finally, using this approach, we detected a novel association between the IL6 receptor signal transduction gene IL6ST and polymyalgia rheumatica, an indication for which sarilumab, a monoclonal antibody against IL-6, has been recently approved. **CONCLUSIONS**: Combining variant annotation and activity-by-contact maps to molQTL increases performance to identify causal genes, while informing on directionality which can be translated to successful target identification and drug development.

## BACKGROUND

Genome-wide associations studies (GWAS) have been successful in identifying genes associated with traits, diseases, and molecular phenotypes.[1, 2] Discoveries from GWAS have increased substantially over the years due to low cost of genomic profiling technologies, an increased number of studies, larger cohorts, and meta-analyses, as well as the formation of deeply phenotyped datasets.[3] The later include large-scale biobank projects such as UK Biobank (UKB)[4, 5], Estonian Biobank[6],and FinnGen.[7] As an example, the UK Biobank alone has contributed to over 3,200 publications (https://www.ukbiobank.ac.uk/enable-your-research/publications), and the FinnGen project is set to increase the number of discoveries emerging from rare variants enriched in the Finnish population.[7] Similarly, the Estonian Biobank, with its extensive dataset, has enhanced rare and low-frequency genetic variation discoveries.[8–10]

Discoveries from genetic studies provide a highly valuable resource for drug discoveries. For example, therapeutic targets with genetic support are >2 times more likely to succeed in clinical trials.[11, 12] A notable example is the association between a loss-of-function missense variant in IL23R gene and Crohn’s disease, suggesting that IL-23 blockage could be beneficial.[13–16] Drugs targeting the IL-23 receptor including Ustekinumab and Risankizumab have recently been approved by the FDA for the treatment of Crohn’s disease following successful clinical trials.[17–19] Other notable examples of targets supported by GWAS include IL6R for rheumatoid arthritis (Sarilumab, Tocilizumab) and HMGCR for high levels of low-density lipoprotein (statins).[20, 21]

While these examples clearly show that disease-associated genetic information is important for drug development, it remains a challenge to accurately assign causal genes driving disease risk from GWAS as most variants identified in GWAS fall in non-coding regions of the genome.[22–24] While it’s been observed that the nearest gene often is the causal gene, this is not a guarantee as genetic variants can influence traits over large genomic distances.[25] In addition, this observation may be biased towards genes that have been well-characterized because they fall at the center of genetic association signals.[26]

Several approaches have been used to predict causal genes, including selecting the nearest gene, variant pathogenicity predictions, epigenetic interactions, and integration of molecular quantitative trait loci (molQTL) such as expression QTL (eQTL). Mendelian randomization (MR) integrating GWAS and molQTL can help identify causal relationships while informing on directionality but may be confounded due to linkage disequilibrium (LD). [27–29] On the other hand, colocalization approaches can be used to detect whether molQTL and GWAS signals share a common causal variant in a specific locus.[30, 31] While colocalization approaches can link genetic variation to changes in gene expression in specific tissue or cell-type contexts, they also tend to be pleiotropic and often impact the expression of multiple genes within the same locus.[26, 32, 33] They can also impact expression across multiple tissues and cell types, decreasing their utility to identify pathogenic cell types.[32, 34, 35] In addition, a large fraction of GWAS loci don’t show eQTL signals, potentially due to the unavailability of data for relevant cell types or specific biological contexts or variants affecting disease risk due to different mechanisms such as splicing.[32, 36, 37] Despite these challenges, eQTL have successfully been used to identify causal genes.[38, 39]. In addition, recent prioritization approaches such as the Locus to Gene (L2G) scores from Open Targets have shown that incorporating molecular trait information does increase performance to identify relevant genes.[26]

Here, we sought to use currently available eQTL information to identify disease relevant genes in the context of drug discovery. We first derived a simple approach to prioritize causal genes based on MR[40], eQTL colocalization[31], activity-by-contact (ABC) enhancer-promoter interactions[41], and variant annotations[42]. We used this combinatorial approach as a way to mitigate the pleiotropic effect of eQTL while retaining important information about directionality. We show that this approach enriches for gold standard genes[26] and captures known target biology. In addition, genes prioritized by this approach are enriched for drug targets with successful clinical trials, and directionality inferred by MR or coding variants recapitulate drug mechanisms of action (MoA). Finally, we show that this approach can be used to identify drug indication expansion opportunities using genes related to the IL6-R as a case study and identify a novel association between IL6ST and polymyalgia rheumatica.

## METHODS

### Estonian Biobank GWAS

The Estonian Biobank (EstBB) is a population-based biobank with 200k participants. The 198k data freeze was used for the analyses described here. All biobank participants have signed a broad informed consent form.

All EstBB participants have been genotyped at the Core Genotyping Lab of the Institute of Genomics, University of Tartu, using Illumina Global Screening Array v1.0 and v2.0. Samples were genotyped and PLINK format files were created using Illumina GenomeStudio v2.0.4. Individuals were excluded from the analysis if their call-rate was <95% or if sex defined based on heterozygosity of X chromosome did not match sex in phenotype data. Before phasing and imputation, variants were filtered by call-rate <95%, HWE p valuel]<l]1e-4 (autosomal variants only), and minor allele frequency <1%. Variant positions were updated to b37 and all variants were changed to be from TOP strand using GSAMD-24v1-0_20011747_A1-b37.strand.RefAlt.zip files from https://www.well.ox.ac.uk/∼wrayner/strand/ webpage. Chip data pre-phasing was done using Eagle v2.3 software [43] (number of conditioning haplotypes Eagle uses when phasing each sample was set to:–Kpbwt=20000) and imputation was done using Beagle v.28 Sep18.7932 [44] with effective population size nel]=l]20,000. Population specific imputation reference panel of 2297 WGS samples was used.[44]

### FinnGen

The FinnGen study (https://www.finngen.fi/en) was described previously.[7] The study is a public-private research project that combines genetic and healthcare data of over 500,000 Finns. The objective of the FinnGen study is to identify novel medical and therapeutical insight into human diseases. It is a pre-competitive partnership of Finnish biobanks, universities and university hospitals, international pharmaceutical industry partners, and Finnish biobank cooperative (FINBB). A full list of FinnGen partners is published here: https://www.finngen.fi/en/partners.

### Disease GWAS processing

We retrieved GWAS results from FinnGen release 10 (R10), UK Biobank pan meta-analysis[45], and a meta-analyses between FinnGen, UK Biobank, and Estonian biobank. For simplicity, we use the term GWAS to refer to both single study GWAS and meta-analyses throughout the manuscript. In total, we included 4,611 GWAS with at least one variant with P<1×10^-6^. When appropriate, we lifted over variants from hg38 to hg19 using the liftOver R package[46]. Variant with a minor allele frequency (MAF) < 0.0001 were excluded from the analysis. For each GWAS, we considered genes located within 250kb of a variant with P<1×10^-6^ for further analysis. For gold standard and clinical trial enrichment analyses (described below), only genome-wide significant loci were included (P<5×10^-8^). We excluded the human leukocyte antigen (HLA) region in all analyses.

### Disease EFO mapping

In order to perform semantic integration of genetic data and clinical trial data, the ontological system Experimental Factor Ontology (EFO) was used. We used the EFO to map traits to their corresponding EFO categories and when multiple EFO terms could be mapped to the same trait, we assigned the trait to each possible term. We used the EFO version 3.52.0 (https://www.ebi.ac.uk/efo/).

### Variant annotation

We used variant effect predictor (VEP v102) [42] to annotate the impact of variants with the following options: ––everything ––offline ––check_existing. Coding variants were defined as those impacting protein coding transcript annotated as missense variant or predicted to have “high” impact. We also retrieved predicted gain or loss of function (GoLoF) variants from LoGoFunc[47], and linked non-coding variants to genes using activity-by-contact (ABC) maps[41]. ABC scores represent the contribution of an enhancer to the regulation of gene, measured by multiplying the estimates of enhancer activity and three-dimensional contact frequencies between enhancers and promoters. ABCmax refers to variant-gene pairs with the highest ABC score. We also retrieved disease mutations from the Human Gene Mutation Database (HGMD) (license acquired via Qiagen, Maryland)[48]

### Mendelian randomization & colocalization

We performed transcriptome wide MR using the R package TwoSampleMR [40]. When more than one instrument was present, we used the inverse variant weighted approach, otherwise we used the Wald Ratio approach. We considered the following exposures: protein quantitative trait loci (pQTL) from Sun et al [49], and expression quantitative trait loci from Blueprint[50], eQTLGen [51] and other datasets from the EBI eQTL catalogue[51–75]. In total, 110 molQTL from 26 studies were included. For each of those studies, we excluded variants with a MAF < 1%. We clumped variants using PLINK[76] using the options –clump-p1 1 –clump-p2 1 –clump-r2 0.01 – clump-kb 10000 and using the European ancestry subset of the 1000 Genomes Project phase 3 data as reference[77]. We only considered genes 250kb around significant loci in this analysis. For each QTL, independent variants with P<1×10^-4^ were used as instruments. For genes with significant MR results (false discovery rate < 0.05), we also performed colocalization analysis using COLOC[31], using a region of 250kb around the local lead GWAS variant. Harmonization between the QTL and GWAS datasets was performed using the harmonise_data function in the TwoSampleMR package[40]. Only autosomes were included in this analysis.

### Causal gene prioritization

We prioritized genes as putatively causal using a combination of evidence including MR, colocalization H4 posterior probabilities (PP) with molQTL, presence of an associated GoLoF variant[47] or other coding variants, distance to lead variant, and enhancer-promoter ABC scores[41]. Specifically, we ranked genes as follow:

| Rank | Criteria |
| --- | --- |
| **Very High** | Lead GoLoF variant;<br>Or<br>Colocalization (H4 PP> 80%) with molQTL of the target gene in >2 dataset; and maximum ABC score for a regulatory element overlapping the lead variant |
| **High** | Lead coding variant;<br>Or<br>Associated ($P<1\times10^{-6}$) GoLoF variant;<br>Or<br>Colocalization (H4 PP> 80%) with molQTL of the target gene in >2 dataset or significant MR with protein QTL (q-value < 0.05); and maximum ABC score for an associated variant overlapping a regulatory element ($P<1\times10^{-6}$) |
| **Moderate** | Colocalization with molQTL of the target gene (H4 PP>80%)<br>Or<br>Significant MR with genome-wide protein QTL (q-value < 0.05)<br>Or<br>Maximum ABC score for an element overlapping the lead variant<br>Or<br>Associated ($P<1\times10^{-6}$) coding variant |
| **Weak** | Colocalization with molQTL of the target gene (H4 PP>30%)<br>Or<br>Nearest gene to the lead variant<br>Or<br>Maximum ABC score for an element overlapping an associated variant ($P<1\times10^{-6}$)<br>Or<br>ABC link (any score) between an element overlapping the lead variant and target gene |
| **Very weak** | Significant MR with eQTL<br>Or<br>ABC link (any score) between an element overlapping the lead variant and target |
|  | gene |

For a given locus, we then prioritized the best gene(s) as the one with the highest rank. In case of ties, we prioritized the nearest gene to lead variant if it is within the set of genes with highest scores, otherwise all highest ranked genes were prioritized equally.

### Enrichment of gold standard genes

We retrieved GWAS causal gene gold standards supported by functional experiments or observations or expert curation from Open Targets (version 191108).[26, 78] We linked the current analysis with the gold standard gene list using Ensembl gene identifiers and EFO codes. That is, for a given gene-disease pair in the current analysis, we consider it a gold standard association if the gene and GWAS EFO code are present in the Open Targets gold standard gene-disease set. For each indication, we filtered out genes not represented in loci where a gold standard gene is located. We calculated the enrichment of gold standard genes in prioritized genes by different features or rankings as described above using Fisher exact tests. In addition, we calculated the precision (number of prioritized genes that are gold standards over all prioritized genes), recall (number of prioritized genes that are gold standards over the total number of gold standard genes), and F1 scores for each feature.

### Single gene colocalizing cell-type eQTL enrichment

To identify enriched colocalizing cell types for single genes, we calculated the ratio of indications for which this gene is prioritized to be causal by a given molQTL dataset (H4 PP > 80%) over the total number of prioritized indications (as defined by unique EFO) for that gene. We collapsed GWAS by corresponding EFO code so that a gene was only counted once per indication (and not multiple times for GWAS of the same disease). We then compared this ratio to the fraction of prioritized indications via colocalization of the same eQTL dataset over all prioritized indications genome wide. In other words, we are looking for genes that show an overrepresentation of colocalizing eQTL cell types across all associated indications compared to the genome-wide distribution. This corresponds to the following contingency table:

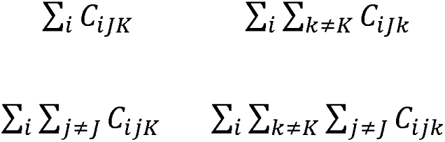

Where C_ijk_=1 if disease i colocalize with prioritized gene j in tissue k and 0 if not. P-values and odds ratios were calculated using Fisher exact tests. False discovery rate (FDR) adjusted P-values < 0.05 were considered significant.

### Enrichment of disease categories for single genes

To identify enrichment disease categories for single genes, we calculated the ratio of the number of GWAS where the genes is prioritized for a given EFO category over the total number of prioritized GWAS for that gene. We then compared this ratio to the genome-wide ratio of GWAS for this EFO category over the total number of tested GWAS. This corresponds to the following contingency table:

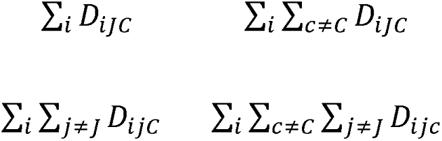

Where D_ijk_=1 if disease i is prioritized for gene j and belongs to category c and 0 if not. P-values and odds ratios were calculated using Fisher exact tests. FDR adjusted P-values < 0.05 were considered significant.

### Disease colocalizing molQTL cell-type enrichment

We identify enriched cell types in GWAS disease EFO categories supported by colocalization as in King et al. 2021.[79] Briefly, we extracted all GWAS colocalizing molQTL (H4 probability > 0.8). Then, for a given cell type K and disease category I, we generated the following contingency table:

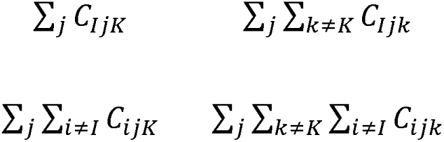

Where C_ijk_=1 if at least one disease GWAS of category i colocalize with gene j in tissue k and 0 if not. P-values and odds ratios were calculated using Fisher exact tests. We performed the analysis considering all molQTL separately, as well as by grouping similar cell types and tissues together prior to testing for enrichment. FDR adjusted P-values < 0.05 were considered significant.

### Drug target-indication pairs in clinical trials

Information about drugs approved or in clinical trials was obtained from the Citeline data from Informa Pharma Intelligence, which is a superset of the most used data sources. In addition to multiple data streams, including nightly feeds from official sources such as ClinicalTrials.gov, Citeline also contains data from primary sources such as institutional press releases, financial reports, study reports, and drug marketing label applications, and secondary sources such as analyst reports by consulting companies. Secondary sources are particularly important to reduce potential biases to the organizations’ tenancy to report only successful trials, especially those before the FDA Amendments Act of 2007, which requires all clinical trials to be registered and tracked by ClinicalTrials.gov. Citeline database contains information from both US and non-US sources. Any cancer or cancer related indications were excluded from this analysis.

In order to map gene-disease pairs in the genetic data to target-indication pairs in the drug data, we used experimental factor ontology (EFO), which provided a systematic description of many data elements available in EBI databases. A target-indication pair is said to have genetic evidence if there is genetic evidence of association between the gene and disease sufficiently similar to the indication, based on semantic similarity. Two methods were used to calculate semantic similarity matrix.[80, 81] Semantic similarities between each pair of EFO headings were computed in the ontologySimilarity R package.[82] The average of the two methods was calculated and standardized similarities had a maximum value of 1 for each disease or indication. Two diseases are considered similar if the similarity is greater than or equal to a previously published value of 0.7.[11]

### Prediction of drug mechanism of action directionality

We retrieved information about drug mechanism of action from the Informa Pharma Intelligence dataset described above. For targets for which decreased expression or loss of function (LoF) is beneficial, we considered datasets with the following keywords: “antagonist”, “inhibitor”, and “degrader”. For targets for which increased expression or function is beneficial, we considered the following keyworks: “agonist”, and “activator”. We considered drugs and targets in phase II clinical trial or above. We performed two analyses to infer directionality from GWAS. First, we assess directionality using the effect size of low-frequency lead coding variant (MAF < 5%). We assumed that these variants are disruptive or LoF. Therefore, a LoF coding variant associated with increased risk suggests that a drug MoA of agonist or activator would be beneficial, whereas for a protective LoF coding variant, an inhibitor or antagonist would be beneficial. Next, we assessed directionality based on the direction of effect of gene expression on disease risk predicted by MR using molQTL as exposure (q-value < 0.05). We included only molQTL colocalizing with local GWAS signal (H4 PP > 80%). For gene-disease pairs supported by multiple colocalizing molQTL, a consensus direction was inferred if the MR direction of effect was consistent across > 75% of the molQTL. Here, a negative consensus MR direction suggests that increased gene expression leads to decreased disease risk. Therefore, an activator or agonist drug targeting this gene would be beneficial. Conversely, a positive consensus MR direction suggests that increased gene expression increases disease risk, and an inhibitor or antagonist drug would be beneficial. We calculated enrichment of concordant direction of effect between GWAS and drug MoA using Fisher exact tests.

### Identification of causal links between diseases and genes related to the IL6 receptor

We aimed to apply our proposed approach to a specific case example. Using the causal gene prioritization and GWAS datasets described above, we extracted all disease GWAS for which IL6, IL6R, or IL6ST were predicted to be causal. We predicted directionality of effect of gene expression on disease risk by MR as above using a threshold of q-value < 0.05. We generated local association of plots molQTL and GWAS using LocusZoom[83]. We performed fine-mapping of IL6ST genetic variants associated with polymyalgia rheumatica using SuSIE[84] as previously described for FinnGen[7].

## RESULTS

### Prioritization of putative causal genes in thousands of GWAS

We aimed to prioritize causal genes across 4,611 GWAS from 3 different sources (Table 1): UK Biobank (UKB)[45], FinnGen release 10 (R10), and meta-analyses of UK Biobank, FinnGen R10, and Estonian biobank.[6] For simplicity, we refer to both single studies and meta-analyses as GWAS throughout the manuscript. While molecular QTLs (molQTL) such as expression quantitative trait loci (eQTL) have been used previously to prioritize causal genes, they are often pleiotropic with the same variant associated with multiple genes within the same locus.[26, 32, 33] Additional genomic information such as the ABC model have been shown to increase performance to identify causal genes, in particular when selecting genes with the highest ABC score (ABCmax).[41] Therefore, we derived a ranking scheme to prioritize genes using different features including ABC, molQTL, presence of an associated coding or gain or loss of function (GoLoF) variants, and distance to lead variant (Figure 1A, methods). We integrated 110 molQTL datasets from 26 studies using MR to infer causality and directionality of gene expression on disease risk. We also performed colocalization analysis to confirm that both GWAS or meta-analyses and molQTL signals shared at least one causal variant. Top ranking genes were selected as those that either contained an associated lead coding variant or were supported by both ABCmax and colocalization across >2 cell types or tissues. We did not include distance to lead variant for higher ranks because we wanted to first prioritize genes for which we could identify potential biological mechanisms. However, for loci without such evidence, or in cases where multiple genes showed identical ranks, the nearest gene to the lead variant was selected as the putative causal gene if it was among the best candidates. Overall, between 1.1 and 1.4 genes were prioritized per locus (before breaking ties with the nearest gene), with 17-49% of loci supported by molQTL colocalization or coding variants (Table 1).

**Figure 1.**
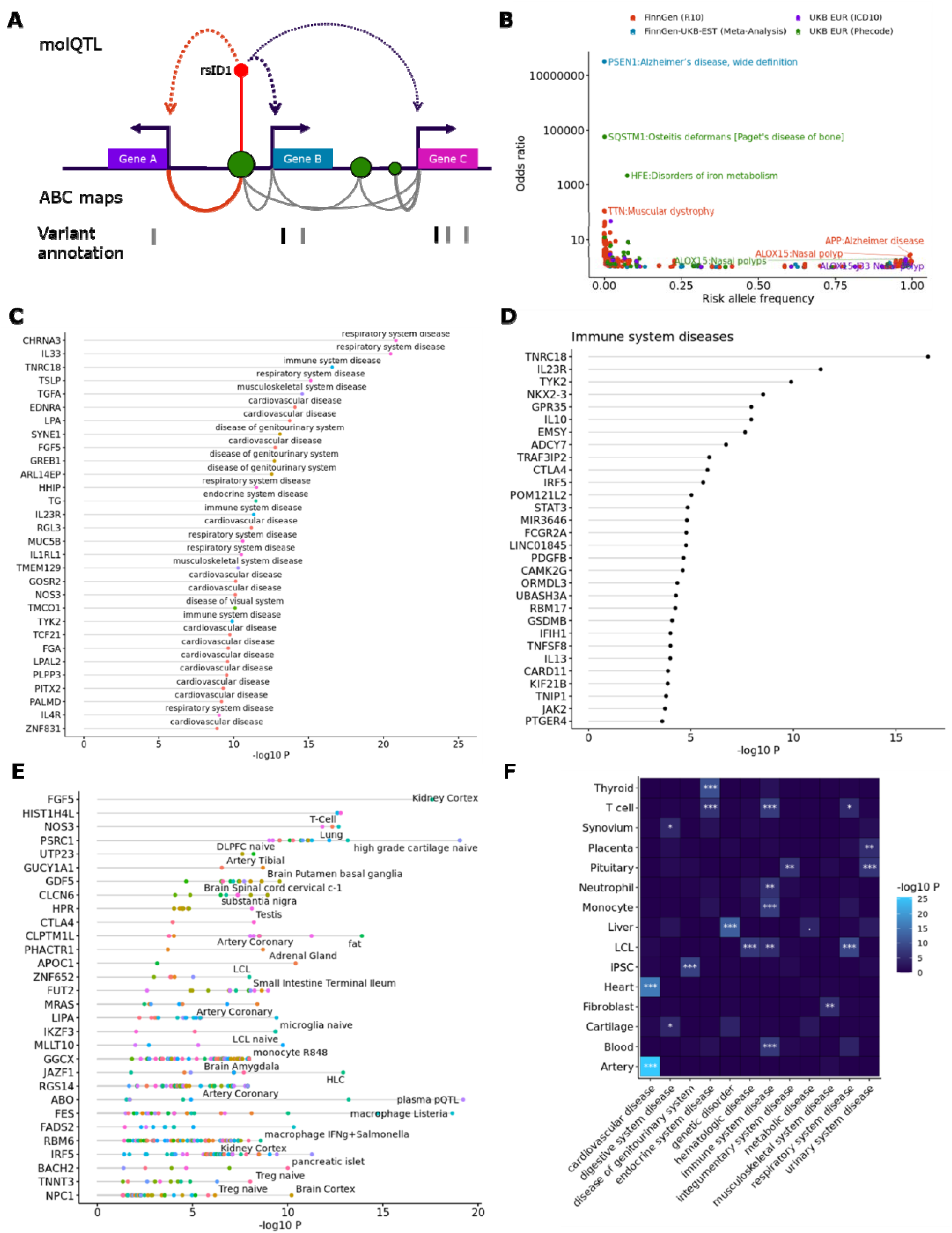
**Characteristics of prioritized genes via gain or loss of function variants and molQTLs**. **A**) Features used to prioritize genes in GWAS loci. Genes are ranked based on a combination of features including molQTLs, activity-by-contact (ABC) maps, and variant annotations, including variant effect predictions (VEP) and loss-of-function (LoF) and gain-of-function (GoF) predictions. **B**) Disease-associated predicted GoF and LoF variants captures disease associations with high effect sizes. Lead GoF and LoF variant with GWAS P-value < 5×10^-8^ are reported in the figure. Effect of the risk allele (odds ratio) is reported on the y-axis. The x-axis corresponds to the frequency of the risk allele. **C**) Disease category overrepresentation for single genes predicted to be causal. Each dot represents a different associated disease category. Top 30 enrichments are shown. **D**) Same as B, but filtered for genes predicted to be causal and enriched in “Immune system diseases”. Each dot represents a different associated disease category. Top 30 genes are shown. **E**) Overrepresentation of eQTL colocalization for single genes predicted to be causal. Gene-tissue pairs are included only if the gene has the highest rank in a locus for a given associated disease. Top 30 colocalized eQTLs are shown. Each dot represents a different enriched tissue or cell-type. **F**) Enriched colocalizing cell types and tissues by disease categories. Only disease categories and tissues or cell types with at least one significant enrichment are reported in the heatmap. Enrichment P-values are calculated using Fisher exact test, testing for the enrichment of genes with eQTL colocalizing with GWAS belonging to specific disease categories as in [79]. Tissues and cell-types were collapsed into broader categories before testing for enrichment. For example, tibial, coronary, and aorta arteries were grouped into “artery”. molQTL: Molecular QTL; ABC: Activity-By-Contact; LCL: Lymphoblastoid cell lines; iPSC: induced Pluripotent Stem Cells.: Adjusted P<0.1; *: Adjusted P<0.05; **: Adjusted P<0.01; ***: Adjusted P<0.001

**Table 1.**
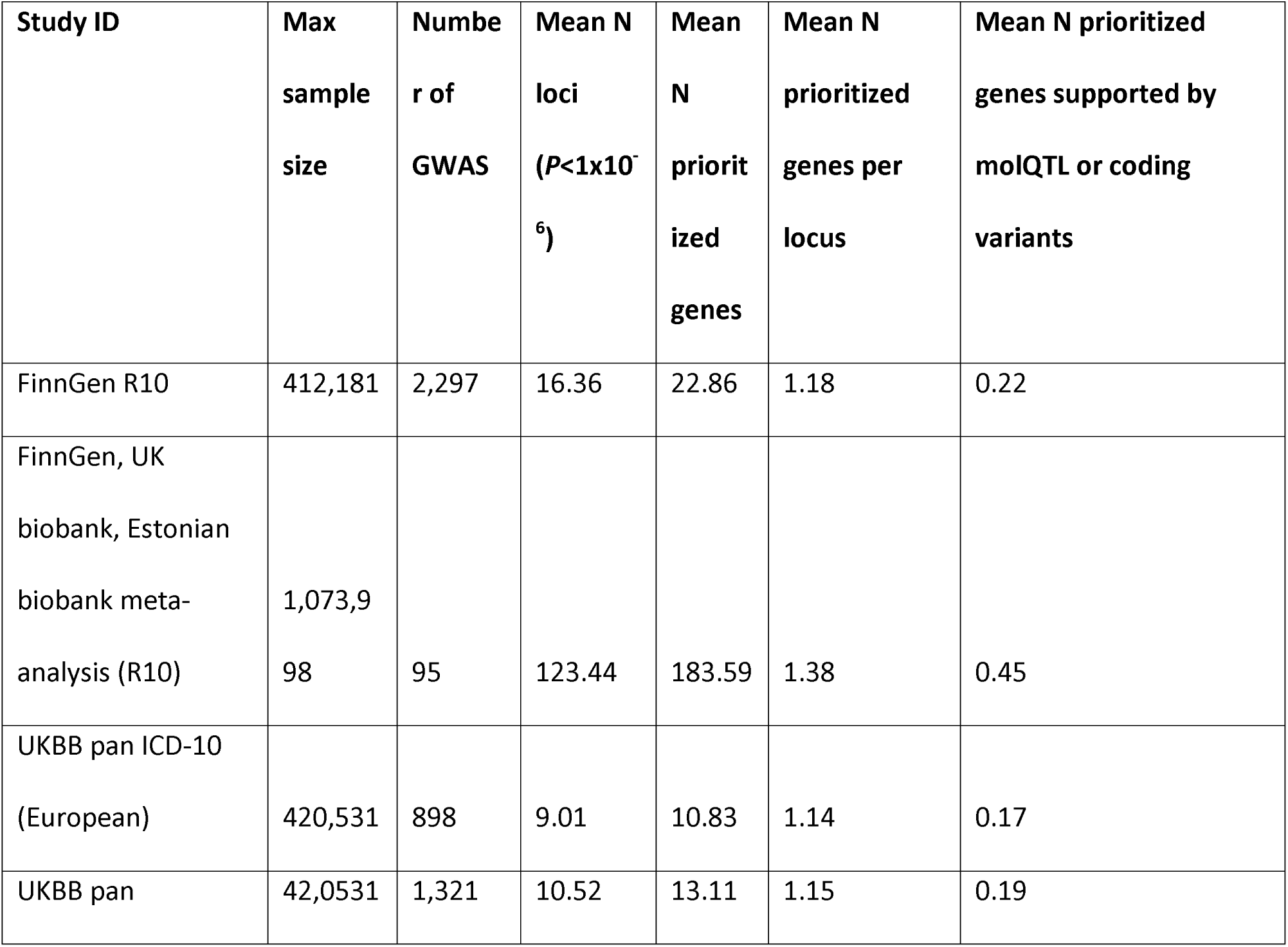
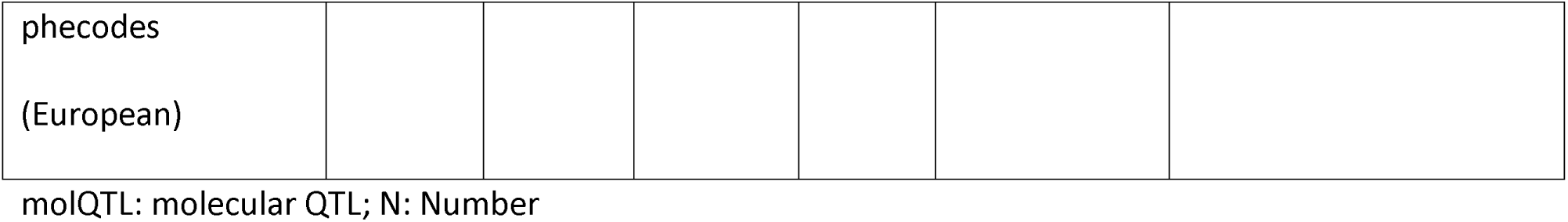
GWAS included in this study. The table reports the maximum GWAS sample size for each study, the total number of GWAS with at least one associated gene. The number of loci with at least one variant with GWAS P<1×10^-6^ To calculate the number of loci, we defined 250kb regions each side of the lead variant. Overlapping regions were then merged. The table reports the total number of non-overlapping regions. The mean number of prioritized genes corresponds to the average number of genes prioritized across each GWAS. The mean number of prioritized gene per locus correspond to the average number of genes with the highest scores in a locus. For the analyses reported throughout this manuscript, ties are broken using the shortest distance to the lead variant. Finally, the last column reports the average number of prioritized gene supported by coding variants or molQTL colocalization.

### Enrichment of genomic features for gold standard genes

Comparing the enrichment of different genomic features alone for curated gold standard genes[26], we found a strong enrichment for genes supported by ABCmax with lead variant (Odds ratio (OR)=8.0-18.7, P=0.0002-4×10^-6^) (Additional file 1: Figure S1; Additional file 2: Table S1). molQTL colocalization also enriched for gold standard genes (colocalization H4 posterior probability (PP) > 95%, OR=3.4-17.7, P=0.001-2×10^-12^). However, the strongest enrichment was generally observed for genes with associated lead coding variants[47] (OR>36.2, P=0.0002-2×10^-10^) and the nearest gene (OR=17.7-38.7, P=3×10^-9^-1×10^-25^). The strong enrichment for nearest genes is expected given that the gene closest to the lead variant is often the causal gene. In addition, several of the gold standard genes have been selected because they are supported by coding variants or tend to fall in the center of GWAS peaks and have been investigated more closely[26]. However, when using these features in combination, we found that our ranking approach performed well and generally better than selecting the nearest gene alone, with a mean increase in F1 score of 0.08 (–0.03 – 0.23) (Additional file 1: Figure S2-S3; Additional file 2: Table S1).

### Gain and Loss of function variants identify genes linked to monogenic disorders

Integrating information about GoLoF variants retrieved variants linked to monogenic disorders including PSEN1 with Alzheimer’s disease (AD)[85] (rs764971634, p.Ile437Val, P=2×10^-12^), SQSTM1 and Paget’s disease[86] (rs104893941, p.Pro392Leu, P=6×10^-11^), and HFE and disorders of iron metabolism[87] (rs1800562, p.Cys282Tyr, P=1×10^-178^) (Figure 1B; Additional file 2: Table S3). We also identified protective GoLoF variants such as APP p.Ala673Thr (rs63750847, P=7×10^-11^) reducing odds of developing AD[88], and ALOX15 p.Thr560Met protecting against nasal polyps (rs34210653, P=2×10^-15^)[89]. Of 208 genes prioritized with at least one predicted GoLoF variant, 179 had at least one disease mutation reported in the Human Gene Mutation Database (HGMD)[48] (OR = 2.3 [1.5-3.6], P=5×10^-6^). Potential novel associations included COLGALT2 and arthrosis (rs35937944, p.Tyr212Cys, P=2×10^-14^), LRG5 and carcinoid syndrome (rs200138614, p.Cys712Phe, P=4×10^-9^), and GREB1 and female infertility (rs755857714, p.Arg1339His, P=4×10^-9^).

### Colocalizing molQTL link genes to diseases and pathogenic tissues

Prioritized candidate causal genes showed enrichment in disease colocalizing molQTLs related to their known function. For instance, colocalizing molQTL for prioritized genes supported associations with disease categories such as EDNRA, LPA and FGF5 with cardiovascular diseases (P<2×10^-16^), TSLP, IL33 and CHRNA3 and respiratory system diseases (P<7×10^-21^), and Il23R, TYK2, IL10 and immune system disease (P<5×10^-11^) (Figure 1C-D; Additional file 2: Table S4). In addition, we found an enrichment of disease colocalizing eQTLs in kidney cortex for FGF5, a gene expressed during kidney development and associated with kidney function (P=4×10^-15^)[90] (Figure 1E; Additional file 2: Table S5). Other examples include artery eQTLs for the cardiovascular diseases associated gene PHACTR1[91] (P=1×10^-9^); the lysosomal acid lipase (LIPA) gene and microglia eQTLs (P=1×10^-10^); and the ABO with plasma pQTL (P=1×10^-20^). Finally, we confirmed that enriched colocalizing eQTLs matched the expected pathogenic tissues and cell-types of different disease categories (Figure 1F; Additional file 2: Table S6). For instance, after grouping eQTL of similar tissues and cell types together, we found a strong enrichment of genes with artery and heart eQTL colocalizing with cardiovascular disease GWAS (P< 9<x10^-17^). We found similar enrichment for T cell and thyroid eQTLs in endocrine system diseases (P<3×10^-8^); blood, lymphoblastoid cell line, monocytes, neutrophil, and T cells with immune system diseases (P<4×10^-6^); and fibroblasts and musculoskeletal diseases (P<4×10^-6^). Treating each eQTL data separately revealed additional associations with tissues or cell subsets including brain cortex and diseases of the visual system (P<6×10^-6^); cerebellum and nervous system diseases (P<4×10^-6^); regulatory T cells and endocrine system diseases (P<9×10^-9^); and T helper 17 cells and digestive system diseases (P<5×10^-7^) (Additional file 1: Figure S4; Additional file 2: Table S7). Overall, the analyses illustrate that in contrast to the nearest gene approach, inclusion of eQTL can help identify potential pathogenic cell types and tissues.

### Prioritized genes increase clinical trial probability of success

Building on these results, we tested whether we could use molQTL information of putative causal gene to drive drug repurposing opportunities or identify potential safety concerns. First, we evaluated whether the prioritized genes enriched for therapeutic targets with clinical trial success. Clinical trial information was retrieved from the Citeline Pharma Intelligence project. Consistent with previous observations, we found that targets with clinical trial success were enriched for features such as presence of coding variation (Figure 2A, Additional file 2: Table S8). For example, gain or loss of function lead variants demonstrated some of the best predictive performances, in particular using genetic evidence from the UKB EUR ICD10 (Phase I: Risk ratio (RR)=1.23, P=0.104; Phase II: RR=1.33, P=0.0688; Phase III: RR=2.08, P=0.0023; Approved: RR=2.67, P=0.00378). Similar results were observed across all studies. Use of epigenetic evidence also improved predictions, for example, lead SNPs linked by the ABC model in UKB EUR ICD10 (Phase I: RR=1.33, P=0.00484; Phase II: RR=1.4, P=0.0162; Phase III: RR=2.15, P=0.000304; Approved: RR=2.82, P=0.000622). However, molQTL information alone did not enrich as much for clinical trial success, for example, colocalizing molQTL with posterior probability > 80% in UKB EUR ICD10 (Phase I: RR=1.22, P=0.013; Phase II: RR=1.18, P=0.154; Phase III: RR=1.43, P=0.0581; Approved: RR=1.71, P=0.044). While the overall prioritized genes did not show the strongest enrichment (UKB ICD10 Phase I: RR=1.24, P=0.0006; Phase II: RR=1.17, P=0.0.08; Phase III: RR=1.51, P=0.003; Approved: RR=1.60, P=0.03), this was likely due to the inclusion of genes with no supportive evidence other than distance (Figure 2A). Indeed, we found that “High” and “Very High” prioritization ranks were more predictive of successful clinical trial progression (higher risk ratios) than lower-ranking genes, especially at later clinical trial phases or approval (High + Very high ranks in UKB ICD10 Phase I: RR=1.16, P=0.103; Phase II: RR=1.18, P=0.174; Phase III: RR=1.78, P=0.00149; Approved: RR=2.06, P=0.00637) (Figure 2B; Additional file 2: Table S9). In our analysis, distance itself was seldom predictive or clinical trial success (UKB ICD10 Phase I: RR=1.18, P=0.03; Phase II: RR=1.06, P=0.0.61; Phase III: RR=1.24, P=0.61; Approved: RR=1.38, P=0.19) especially after excluding loci potentially driven by coding variants (Figure 2B).

**Figure 2.**
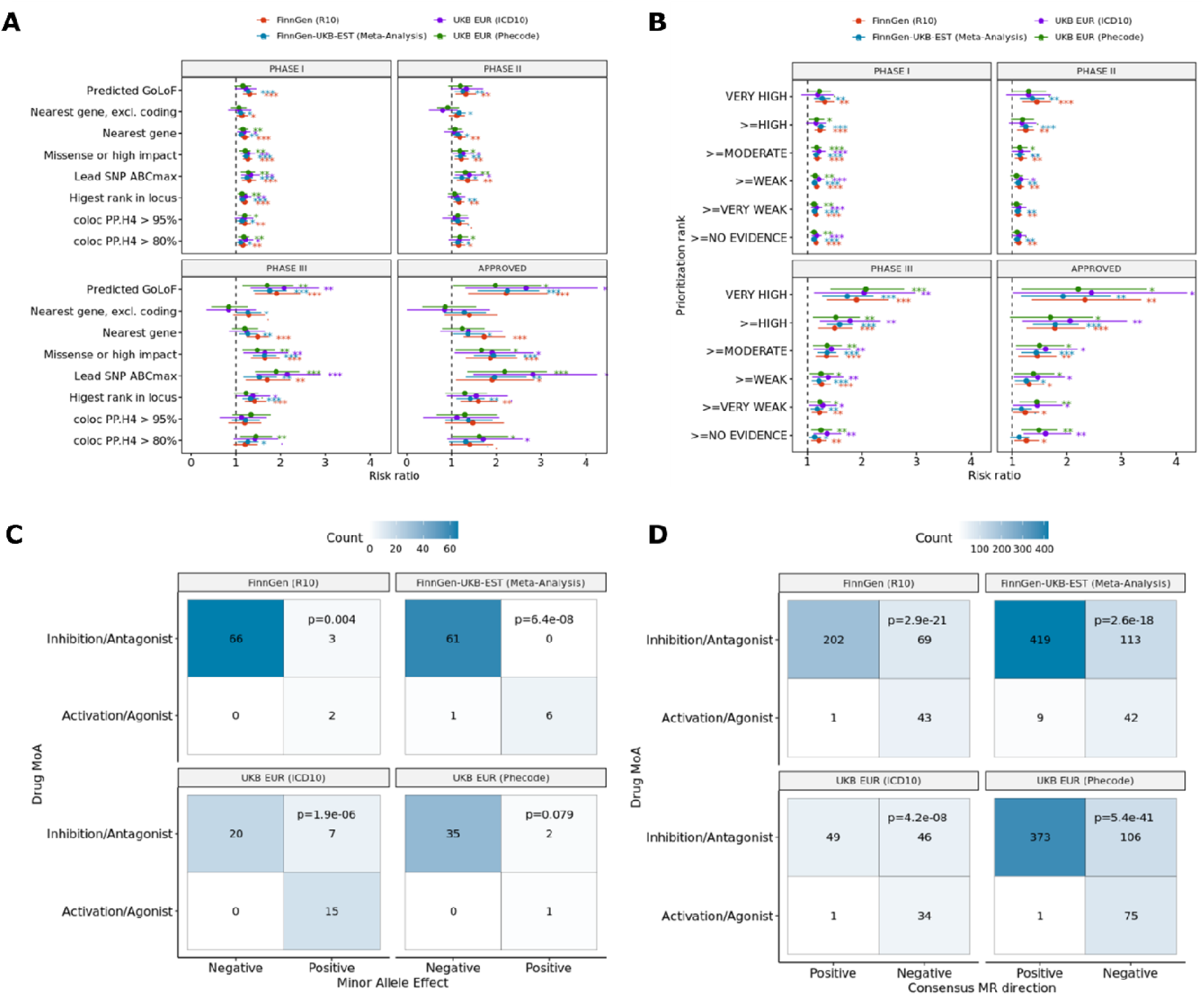
**Prioritized genes predict clinical trial success**. Enrichment of targets of approved drugs or drugs in clinical trials (phase I-III) using genetic evidence from FinnGen, UK Biobank, and biobank meta-analyses prioritizing genes using colocalization (posterior probability of colocalization [H4] > 80% or > 95%), predicted gain of function (GoF) or loss of function (LoF) variants[47], genes with highest prioritization rank, ABC score for lead variant, or nearest gene excluding loci with associated coding variants. **B**) Enrichment of targets of approved drugs or drugs in clinical trials (phase I-III) using causal gene prioritization ranks in FinnGen, UK Biobank, and biobank meta-analyses. **C**) Concordance between direction of effect of lead low-frequency coding variants on disease risk, and drug mechanism of action (MoA) for targets in phase II clinical trials or above. We retrieved information about targets, clinical trials, and drug MoA from the Citeline Pharmacogenomics dataset. We connected this dataset to GWAS phenotypes using EFO codes and a semantic similarity score > 0.7. We assume that low-frequency coding variants (minor allele frequency < 5%) are disruptive (LoF). Therefore a negative (protective) direction of effect would translate into inhibition or antagonism being beneficial (and vice-versa). **D**) Concordance between the predicted impact of gene expression on disease risk predicted by mendelian randomization (MR), and drug MoA for targets in phase II clinical trials or above. Information about targets, clinical trials, and drug MoA were collected from the Citeline Pharmacogenomics dataset and connected to GWAS phenotypes using EFO codes and a semantic similarity score > 0.7. The direction of effect of gene expression on disease risk was assessed by MR using molQTL as exposure (q-value < 0.05). Only molQTL colocalizing with local GWAS signal (H4 posterior probability > 80%) were included. A consensus direction was inferred if the MR direction of effect was consistent across > 75% of molQTL for a given gene and disease GWAS. A negative consensus MR direction suggests that increased gene expression leads to decreased disease risk. Therefore, an activator or agonist drug targeting this gene would be beneficial. Conversely, a positive consensus MR direction suggests that increased gene expression increases disease risk, and an inhibitor or antagonist drug would be beneficial. Reported P-values were calculated by Fisher exact test. .: P<0.1; *: P<0.05; **: P<0.01; ***: P<0.001

### Inferred directionality from GWAS recapitulate drug mechanisms of action

To understand whether inferred directionality could inform on clinical trial success, we first investigated the consistency between the direction of effect of coding variants and drug mechanism of action (MoA) (methods). When considering prioritized genes with lead low-frequency coding variants (minor allele frequency < 0.05) and clinical trials phase II and above, between 83% and 96% of showed consistent effect between the minor allele and drug MoA (Fisher P=0.08-6×10^-8^, Figure 2C). We then asked whether molQTL could similarly inform on directionality. Using prioritized gene-disease pairs supported by MR (q-value < 0.05) and colocalization (PP > 80%), we inferred the direction of effect when the predicted MR effect was consistent across >75% of molQTL datasets for a given gene. This was the case for most gene-disease pairs (Additional file 1: Figure S5). Again, direction of effect was generally in agreement with drug MoA (64-81% agreement, Fisher P=4×10^-8^-5×10^-41^, Figure 2D). Consistency increased when considering only approved drugs (78-93% agreement, Fisher P=3×10^-5^-1×10^-23^, Additional file 1: Figure S6). Overall, these data suggest that molQTL can be used to inform on drug MoA.

### Causal gene predication from GWAS identifies a link between IL6ST and polymyalgia rheumatica

Finally, we applied our causal gene prioritization approach to a specific use case, that is identifying potential new indications for drugs targeting the IL6 receptor such as Sarilumab and Tocilizumab, both drugs approved for rheumatoid arthritis. We extracted diseases prioritized by our approach for genes related to the receptor, namely IL6, IL6ST, and IL6R. We identified putative causal links between increased IL6 expression in CD16 monocytes and increased risk of varicose veins, ischemic heart disease, coronary atherosclerosis, and atrial fibrillation (MR beta > 0), but decreased risk of asthma and allergy (MR beta < 0) (Additional file 1: Figure S7; Additional file 2: Table S10). eQTL of IL6 in whole blood also supported these disease associations, albeit with an opposite predicted direction of effect. Similarly, IL6R expression in multiple tissues including artery, colon, and esophagus was associated with increased risk of coronary revascularization, coronary atherosclerosis, and abdominal aortic aneurysm (AAA), but lower risk of lower respiratory diseases and atopic dermatitis. Again, we observed opposite direction of effect predicted by MR using monocyte or macrophage eQTL as exposure. The associations with coronary atherosclerosis and AAA were further driven by a lead coding variant in IL6R, rs2228145 (Asp358Ala, Additional file 2: Table S10). Finally, we found that increased IL6ST expression in T cells and whole blood is predicted to increase the risk of rheumatoid arthritis, systemic connective tissue disorders, polyarthropathies, other arthritis, autoimmune diseases, and polymyalgia rheumatica (Figure 3A). The later association has not been reported previously to our knowledge. These associations were driven by rs7731626 (SuSIE fine-mapping probability >0.99). This variant is located within an intron of ANKRD55 and colocalizes with eQTLs for both ANKRD55 and IL6ST (PP > 80%). However, this variant also overlaps an enhancer that shows highest ABC score for IL6ST for genes in the region, suggesting the latter is the causal gene, in line with previous studies[92, 93] (Figure 3B). Overall, our approach was able to capture known associations with IL6-R related genes and identified a new association between IL6ST and polymyalgia rheumatica.

**Figure 3.**
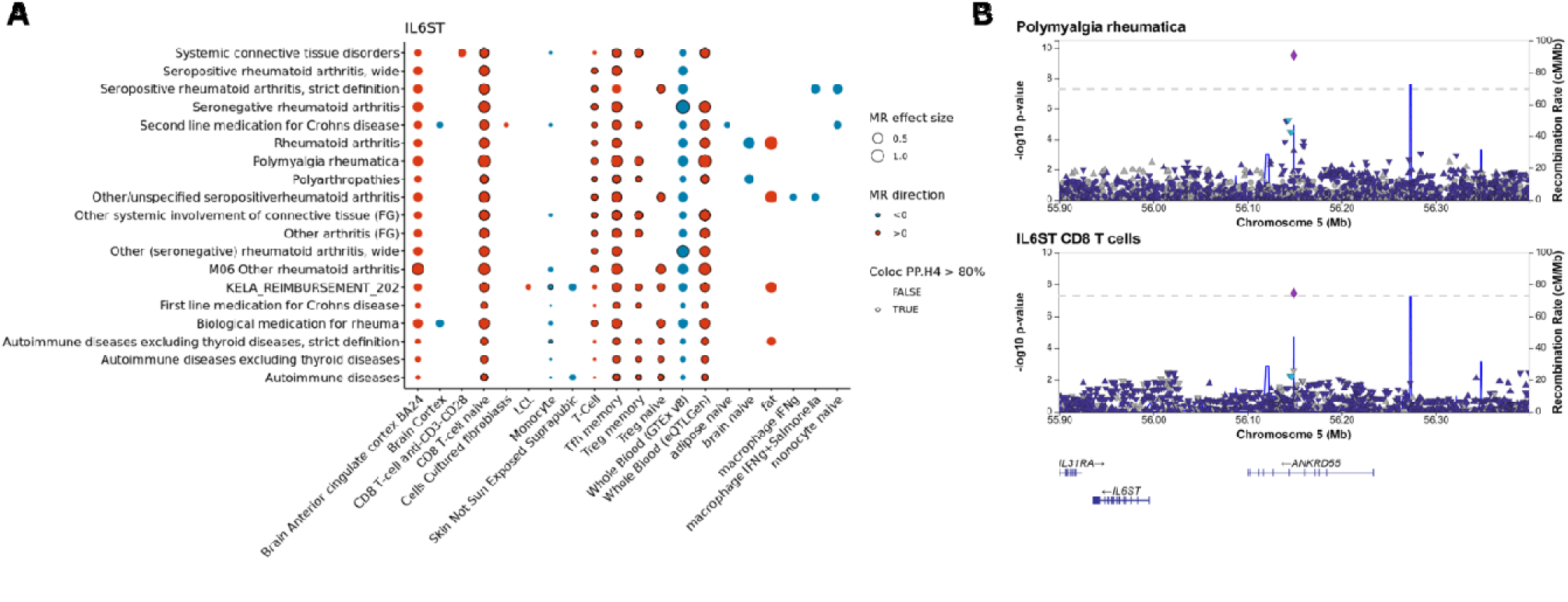
**IL6ST is predicted to be causal for rheumatoid arthritis and polymyalgia rheumatica**. **A**) Diseases associations supported by MR, colocalization and ABC. thows tissues and cell-types with significant MR (q-value < 0.05) using IL6ST eQTL as exposure and diseases as outcome (red: positive effect size estimate [MR beta]; blue: negative effect size estimate). The size of the dots represents absolute effect size. Disease-eQTL pairs with a colocalization posterior probability > 80% are highlighted with a dark border. **B**) LocusZoom[83] plot showing the top association for polymyalgia rheumatica at the ANKRD55-IL6ST locus. Both IL6ST and ANKRD55 eQTL colocalize with the polymyalgia rheumatica signal, but IL6ST has the highest ABC score.

## DISCUSSION

We prioritized disease-associated genes across 4,611 GWAS and meta-analyses from biobank studies using a combination of MR with molQTL, colocalization analysis, variant effect prediction, and epigenetic annotations (ABC model). This approach allows the use of molQTL to infer directionality of gene expression on disease risk, while improving the causal gene prediction compared to using molQTL alone. Based on combination of these features, we used a ranking approach to prioritize genes within loci and showed that this approach enriched for gold standard genes. We recover known coding variant associations, including rare variants in genes linked to monogenic disorders such as PSEN1 and APP1 and Alzheimer’s disease, and SQSTIM1 and Paget’s disease (Figure 1B). Genes prioritized by molQTL also show enrichment in disease categories related to their function with pathogenic tissue contexts (Figure 1C-F). Of note, when multiple genes show evidence of colocalization within the same locus, the addition of epigenetic (ABCmax) information can help prioritize one gene over the others. We note as an example the association of variants with polymyalgia rheumatica at the ANRKD55 locus where this gene would be prioritized using the nearest gene approach. Whereas colocalization alone did not identify a single causal gene, combination of colocalization and ABCmax identified IL6ST as the putative causal gene. To our knowledge, this is the first report of a GWAS association between IL6ST and polymyalgia rheumatica. IL6ST encodes a protein involved in signal transduction for the IL6 receptor pathway. Inhibitors of the IL6 receptor have recently shown success in clinical trials for this indication leading to a recent approval by the FDA.[94]

In line with previous studies[11, 12], we show that therapeutic targets with genetic evidence are enriched at later clinical trial phases and as targets of approved drugs. In our analysis, using the nearest gene information alone was not strongly predictive of clinical trial success. The most predictive features were coding variant annotations and ABC maps. While the later performs well to link causal genes to diseases, it does not provide information about directionality. We used coding variants and MR with molQTL to infer directionality of a target on disease risk. Both approaches were generally consistent with drug MoA matched for the target and disease. These data support that molQTL can be used to predict drug MoA. However, while we found that in general eQTL were consistent across cell type and tissues for a given gene and disease (Additional file 1: Figure S5), we note that this isn’t always the case. This is exemplified by the IL6-R case study, where all three queried gene displayed inconsistent direction of effect predicted by MR depending on the molQTL dataset. Future improvement of this approach should consider prior knowledge on pathogenic cell types or tissues to infer directionality in relevant contexts. Overall, our analysis suggests that using features such as ABCmax in combination to molQTL can increase the performance of causal gene inference approaches while informing on directionality which is crucial for translating GWAS hits to therapies.

We note that this study has some limitations. First, we did not perform fine-mapping analyses nor colocalization approaches that use linkage disequilibrium references. Indeed, we opted to avoid methods that do not rely on LD references as we used GWAS from various sources, including meta-analyses where these methods may not be well calibrated.[95] Nevertheless, using fine-mapping information likely would improve performance, especially in cases where there are multiple causal variants underlying molQTL or GWAS signals, and would reduce LD contamination[30, 96]. In addition, we performed MR and colocalization analyses as separate steps. Tools that use a combination of these approaches have been recently developed, which are likely to perform better in case of allelic heterogeneity[97]. This is evident in the case of IL6ST, where MR using eQTL from whole blood from different sources (GTEx, eQTLGen) lead to inversed estimate of directionality (Figure 3A). This difference was due to different instrument used as only one genetic instrument was included in GTEx whereas 5 independent instruments were included for eQTLGen. We also assume that there is one causal gene per locus, although it is possible that multiple genes contribute to disease risk. Finally, integrating other sources of molQTL such as metabolite or splice QTL could help identify putative causal genes as coding variants and eQTL only cover a fraction of loci (18-45% in this study).[98] While these approaches can be useful to nominate candidate causal genes and their relationship to diseases, proper functional validation remains of high importance.

## CONCLUSIONS

We nominated putative causal genes across 4,611 GWAS from biobank studies and public resources by integrating variant annotations as well as molecular QTL. We show that these prioritized genes recover known biological relationships in terms of disease and tissue enrichment and are enriched for therapeutic targets that succeeded in clinical trials. We show that directionality predicted by molQTL and coding variants generally recapitulate drug mechanism of actions. Finally, we applied this approach to genes related to the IL6 receptor and identified a novel association between IL6ST and polymyalgia rheumatica supporting the recent approval of Sarilumab for this indication.

## ABBREVIATIONS

AAA: abdominal aortic aneurysm; ABC: Activity-by-contact; CI: Confidence interval; EFO: Experimental factor ontology; eQTL: Expression quantitative trait loci; EstBB: Estonian Biobank; GWAS: Genome-wide association study; GoF: Gain of function; GoLoF: Gain or loss of function; HLA: Human leukocyte antigen; iPSC: Induced Pluripotent Stem Cells; LCL: Lymphoblastoid cell lines; LD: Linkage disequilibrium; LoF: Loss of function; MAF: Minor allele frequency; MoA: Mechanism of action; MR: Mendelian randomization; molQTL: Molecular quantitative trait loci; OR: Odds ratio; pQTL: Protein quantitative trait loci; PP: posterior probability; QTL: Quantitative trait loci; RR: Risk ratio; UKB: UK Biobank; VEP: Variant effect predictor.

## DECLARATIONS

### Ethics approval and consent to participate

Patients and control subjects in FinnGen provided informed consent for biobank research, based on the Finnish Biobank Act. Alternatively, separate research cohorts, collected prior the Finnish Biobank Act came into effect (in September 2013) and start of FinnGen (August 2017), were collected based on study-specific consents and later transferred to the Finnish biobanks after approval by Fimea, the National Supervisory Authority for Welfare and Health. Recruitment protocols followed the biobank protocols approved by Fimea. The Coordinating Ethics Committee of the Hospital District of Helsinki and Uusimaa (HUS) approved the FinnGen study protocol Nr HUS/990/2017.

The FinnGen study is approved by Finnish Institute for Health and Welfare (permit numbers: THL/2031/6.02.00/2017, THL/1101/5.05.00/2017, THL/341/6.02.00/2018, THL/2222/6.02.00/2018, THL/283/6.02.00/2019, THL/1721/5.05.00/2019, THL/1524/5.05.00/2020, and THL/2364/14.02/2020), Digital and population data service agency (permit numbers: VRK43431/2017-3, VRK/6909/2018-3, VRK/4415/2019-3), the Social Insurance Institution (permit numbers: KELA 58/522/2017, KELA 131/522/2018, KELA 70/522/2019, KELA 98/522/2019, KELA 138/522/2019, KELA 2/522/2020, KELA 16/522/2020 and Statistics Finland (permit numbers: TK-53-1041-17 and TK-53-90-20).

The Biobank Access Decisions for FinnGen samples and data utilized in FinnGen Data Freeze 6 include: THL Biobank BB2017_55, BB2017_111, BB2018_19, BB_2018_34, BB_2018_67, BB2018_71, BB2019_7, BB2019_8, BB2019_26, BB2020_1, Finnish Red Cross Blood Service Biobank 7.12.2017, Helsinki Biobank HUS/359/2017, Auria Biobank AB17-5154, Biobank Borealis of Northern Finland_2017_1013, Biobank of Eastern Finland 1186/2018, Finnish Clinical Biobank Tampere MH0004, Central Finland Biobank 1-2017, and Terveystalo Biobank STB 2018001.

UK Biobank has received ethical approval from the NHS National Research Ethics Service North West (approval numbers 11/NW/0382 and 16/NW/0274). All participants provided written informed consent.

Estonian Biobank GWAS and consecutive meta-analyses were carried out under ethical approval permit number 1.1-12/1020 from the Estonian Committee on Bioethics and Human Research (Estonian Ministry of Social Affairs).

### Consent for publication

Not applicable.

### Availability of data and materials

The UK Biobank Pan ancestry GWAS[45] are available through https://pan.ukbb.broadinstitute.org/. FinnGen GWAS[7] are available through https://www.finngen.fi/en/access_results. Processed and formatted eQTL data used in this study are available through the eQTL catalogue [52–75] https://www.ebi.ac.uk/eqtl/. pQTL from Sun et al. 2018[49] are available through http://www.phpc.cam.ac.uk/ceu/proteins/. eQTLGen eQTL[51] are available through https://www.eqtlgen.org/phase1.html. 1000 Genomes project phase 3 data[77] is available through https://www.internationalgenome.org/category/phase-3/. Activity-by-contact maps[41] are available through https://www.engreitzlab.org/resources/. LoGoFunc[47] gain and loss of function predictions are available through https://itanlab.shinyapps.io/goflof/. Datasets supporting the conclusions of this article are included within the article and its additional files. Additional datasets used and/or analysed during the current study are available from the corresponding author on reasonable request.

### Authors’ contributions

SL, MC, and CC performed data analysis, interpreted the results, designed analyses, and are major contributors in writing the manuscript. FinnGen authors defined endpoints, performed GWAS, generated summary statistics, and performed meta-analyses. Estonian Biobank authors defined matching disease endpoints, performed GWAS and generated EstBB summary statistics. MB acquired and formatted data and performed data analysis. SS, KK, EdR, JS, SK, DR designed the study, interpreted results, and contributed to writing the manuscript. All authors read and approved the final manuscript.

## Data Availability

The UK Biobank Pan ancestry GWAS[45] are available through https://pan.ukbb.broadinstitute.org/. FinnGen GWAS are available through https://www.finngen.fi/en/access_results. Processed and formatted eQTL data used in this study are available through the eQTL catalogue https://www.ebi.ac.uk/eqtl/. pQTL from Sun et al. 2018 are available through http://www.phpc.cam.ac.uk/ceu/proteins/. eQTLGen eQTL are available through https://www.eqtlgen.org/phase1.html. 1000 Genomes project phase 3 data is available through https://www.internationalgenome.org/category/phase-3/. Activity-by-contact maps are available through https://www.engreitzlab.org/resources/. LoGoFunc gain and loss of function predictions are available through https://itanlab.shinyapps.io/goflof/. Datasets supporting the conclusions of this article are included within the article and its additional files. Additional datasets used and/or analysed during the current study are available from the corresponding author on reasonable request.

## Acknowledgments

We thank all participants and contributors to the datasets used in this study. We thank Hao He (Sanofi US) for his valuable feedback on the manuscript, and Omar Stradella (Sanofi US) for his support with ontology mapping.

## UK Biobank

This research has been conducted using the UK Biobank Resource (www.ukbiobank.ac.uk), a large-scale biomedical database and research resource containing genetic, lifestyle and health information from 500,000 UK participants. UK Biobank is supported by its founding funders the Wellcome Trust and UK Medical Research Council, as well as the Department of Health, Scottish Government, the Northwest Regional Development Agency, British Heart Foundation and Cancer Research UK. The UK biobank pan-ancestry analysis was conducted under project ID 31063 (https://pan.ukbb.broadinstitute.org).

## FinnGen

The FinnGen project is funded by two grants from Business Finland (HUS 4685/31/2016 and UH 4386/31/2016) and the following industry partners: AbbVie Inc., AstraZeneca UK Ltd, Biogen MA Inc., Celgene Corporation, Celgene International II Sàrl, Genentech Inc., Merck Sharp & Dohme Corp, Pfizer Inc., GlaxoSmithKline Intellectual Property Development Ltd., Sanofi US Services Inc., Maze Therapeutics Inc., Janssen Biotech Inc, and Novartis AG. Following biobanks are acknowledged for delivering biobank samples to FinnGen: Auria Biobank (www.auria.fi/biopankki), THL Biobank (www.thl.fi/biobank), Helsinki Biobank (www.helsinginbiopankki.fi), Biobank Borealis of Northern Finland (https://www.ppshp.fi/Tutkimus-ja-opetus/Biopankki/Pages/Biobank-Borealis-briefly-in-English.aspx), Finnish Clinical Biobank Tampere (www.tays.fi/en-US/Research_and_development/Finnish_Clinical_Biobank_Tampere), Biobank of Eastern Finland (www.ita-suomenbiopankki.fi/en), Central Finland Biobank (www.ksshp.fi/fi-FI/Potilaalle/Biopankki), Finnish Red Cross Blood Service Biobank (www.veripalvelu.fi/verenluovutus/biopankkitoiminta) and Terveystalo Biobank (www.terveystalo.com/fi/Yritystietoa/Terveystalo-Biopankki/Biopankki/). All Finnish Biobanks are members of BBMRI.fi infrastructure (www.bbmri.fi). Finnish Biobank Cooperative – FINBB (https://finbb.fi/) is the coordinator of BBMRI-ERIC operations in Finland.

## Estonian Biobank

Estonian Biobank research was supported by the European Union through Horizon 2020 research and innovation programme under grant no 810645 and through the European Regional Development Fund project no. MOBEC008, by the Estonian Research Council grant PUT (PRG1291, PRG687 and PRG184) and by the European Union through the European Regional Development Fund project no. MOBERA21 (ERA-CVD project DETECT ARRHYTHMIAS, GA no JTC2018-009), Project No. 2014-2020.4.01.15-0012 and Project No. 2014-2020.4.01.16-0125. Computations were performed in the High Performance Computing Center, University of Tartu.

## molQTL datasets

This manuscript makes use of previously published molecular QTL data. Except for pQTL from Sun 2018 and eQTL Gen, formatted summary statistics were obtained from the EBI eQTL catalogue. We wish to thank all participants and contributors to these datasets. We list funding sources of each of the dataset in a supplementary note.

## Estonian Biobank and FinnGen banners

The Estonian Biobank team is composed of: Andres Metspalu, Mari Nelis, Lili Milani, Reedik Mägi, Georgi Hudjashov, and Tõnu Esko (University of Tartu, Tartu, Estonia). FinnGen authors and their institution are listed in a supplementary file.

## ADDITIONAL INFORMATION

**Additional file 1 (docx): Supplementary figures S1-S8**. Figure S1: Gold standard gene enrichment by genomic features. Figure S2: Precision and recall of gold standard genes for different genomic features as well as causal candidate prioritization approach. Figure S3: F1 scores for each considered features and prioritization scheme. Figure S4: Enriched colocalizing cell types and tissues by disease categories. Figure S5: Predicted direction of effect of gene expression on disease risk. Figure S6: Concordance between the predicted effect of gene expression on disease risk by mendelian randomization (MR) and mechanism of action (MoA) of approved drugs. Figure S7: Association between IL6 and diseases, supported by MR, colocalization and ABC. Figure S8: Association between IL6R and diseases, supported by MR, colocalization and ABC.

**Additional file 2 (xlsx): Supplementary tables S1-S10**. Table S1: Enrichment of gold standard genes by feature and GWAS study source. Table S2: Precision and recall of different features to recover gold standard genes. Table S3: Genes with predicted gain or loss of function variants (P<1e-6). Table S4: Genes with overrepresented disease categories of GWAS in which they are prioritized as causal. Table S5: Genes with overrepresented cell-type colocalizing QTL with GWAS in which they are prioritized as causal. Table S6: Significantly enriched colocalizing QTL cell types and tissues in disease GWAS categories, after grouping similar tissues and cell-types together. Table S7: Significantly enriched colocalizing QTL cell types and tissues in disease GWAS categories, treating each eQTL dataset separately. Table S8: Enrichment of prioritized genes by feature across clinical trial phases and approved drugs. Table S9: Enrichment of prioritized genes by rank across clinical trial phases and approved drugs. Table S10: Causal association between diseases and IL6, IL6ST, or IL6R.

**Additional file 3: FinnGen banner authors and affiliations**. List of FinnGen authors and their affiliations.

**Additional file 4: Funding statements for eQTL and pQTL datasets**. Funding statements and references for all eQTL and pQTL datasets used for this manuscript.

