## Additional File 1 for "Leveraging large-scale multi-omics to identify therapeutic targets from genome-wide association studies"

**SUPPLEMENTAL FIGURES**

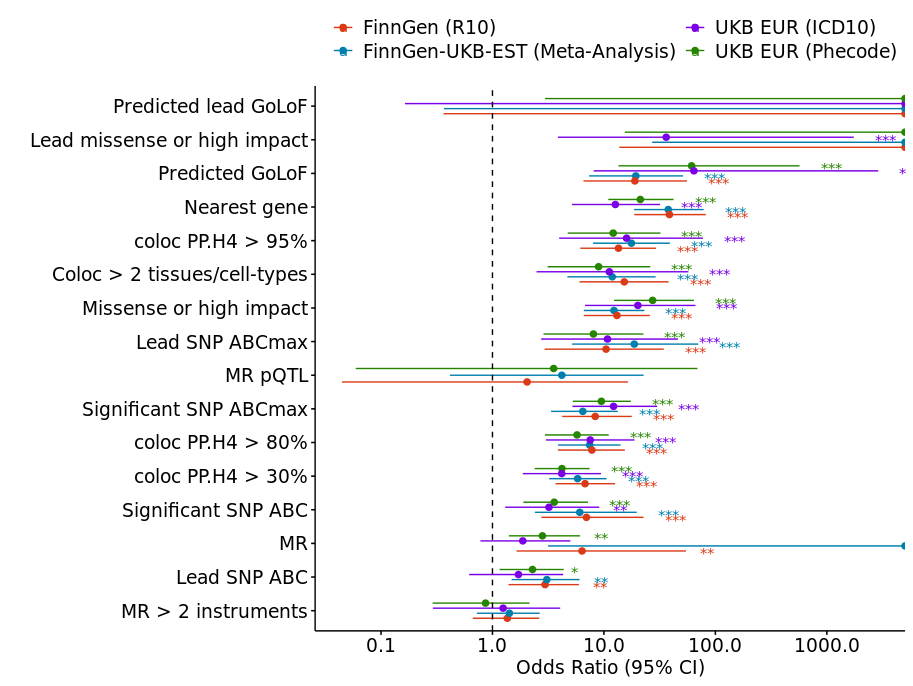

**Figure S1. Gold standard gene enrichment by genomic features.** The figure shows the odds ratio of recovering gold standard genes[1] for each feature separately. For predicted gain or loss of function lead variants (GoLoF) and in some cases of lead missense or high impact variant and mendelian randomization (MR), features recovered all gold standard genes, leading to infinite odds ratio (OR). Predicted gain or loss of function (GoLoF) variants were annotated using a list derived from Stein *et al*.[2] Missense or high impact variant were annotated using variant effect predictor[3]. Nearest genes correspond to gene with their transcription start site nearest to the GWAS lead variant. Coloc > 2 tissues or cell types indicates loci with a colocalization posterior probability (H4) > 80% using eQTL from >2 different cell types or tissues.

GoLoF: Gain or Loss of function; coloc: Colocalization; MR mendelian randomization; ABC: Activity-by-contact; CI: confidence interval.; PP.H4: Posterior probability of colocalization; . : P<0.1; *: P<0.05; **: P<0.01; ***: P<0.001

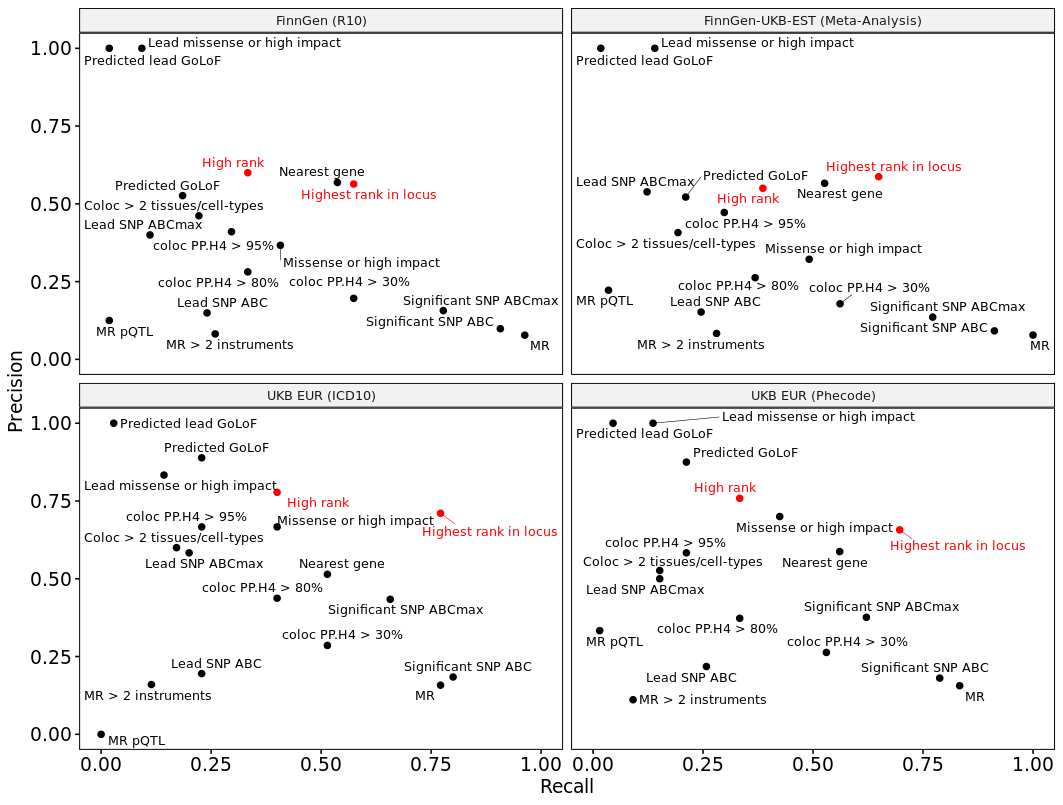

**Figure S2. Precision and recall of gold standard genes for different genomic features as well as causal candidate prioritization approach**. High rank references to genes prioritized for a given disease GWAS locus supported by colocalization >2 cell types and ABC interactions or coding variants (“high” or “very high” ranks, see METHODS). Highest rank in locus refers to genes prioritized for a given disease GWAS locus regardless of rank.

GoLoF: Gain or Loss of function; coloc: Colocalization; MR mendelian randomization; ABC: Activity-by-contact; PP.H4: Posterior probability of colocalization.

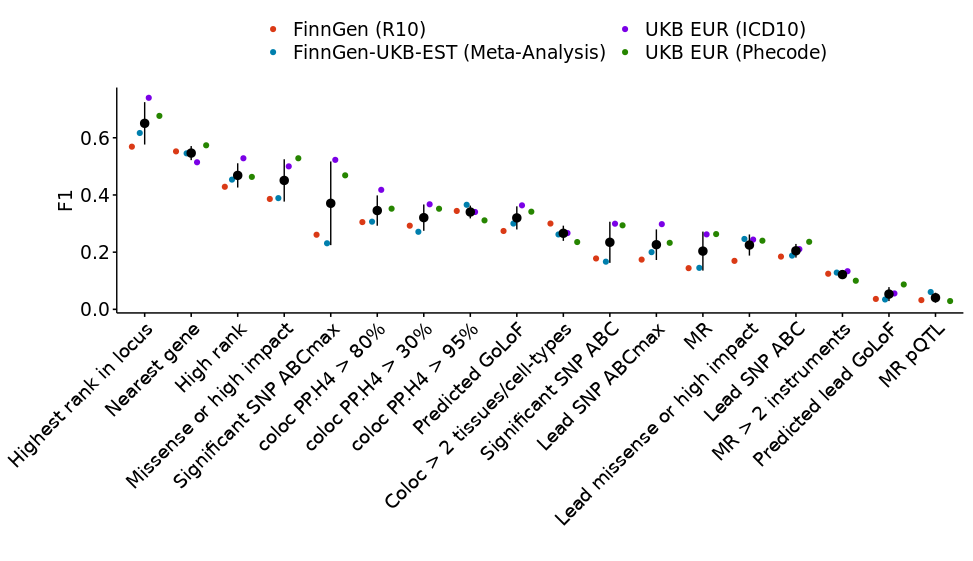
**Figure S3. F1 scores for each considered features and prioritization scheme.** “Highest rank in locus” corresponds to the best scoring gene(s) for a given GWAS within a particular locus. “High rank” corresponds to genes with “high” or “very high” ranks, that is genes supported by an associated coding variant or both ABC and colocalization > 2 cell types or tissues.

GoLoF: Gain or Loss of function; coloc: Colocalization; MR mendelian randomization; ABC: Activity-by-contact; AUC: Area under the curve.

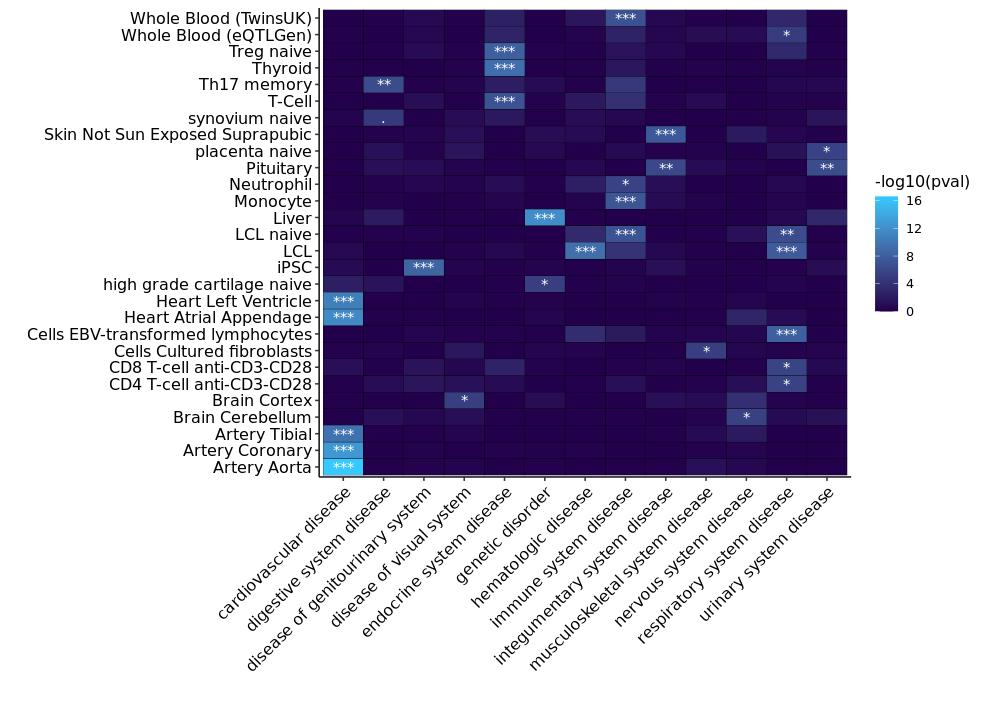

**Figure S4**. **Enriched colocalizing cell types and tissues by disease categories.** Only disease categories and tissues or cell types with at least one significant enrichment are reported in the heatmap. Enrichment *P*-values are calculated using Fisher exact test, testing for the enrichment of genes with eQTL colocalizing with GWAS belonging to specific disease categories as in [4].

Adjusted P<0.1; *: Adjusted P<0.05; ** Adjusted P<0.01; *** Adjusted P<0.001

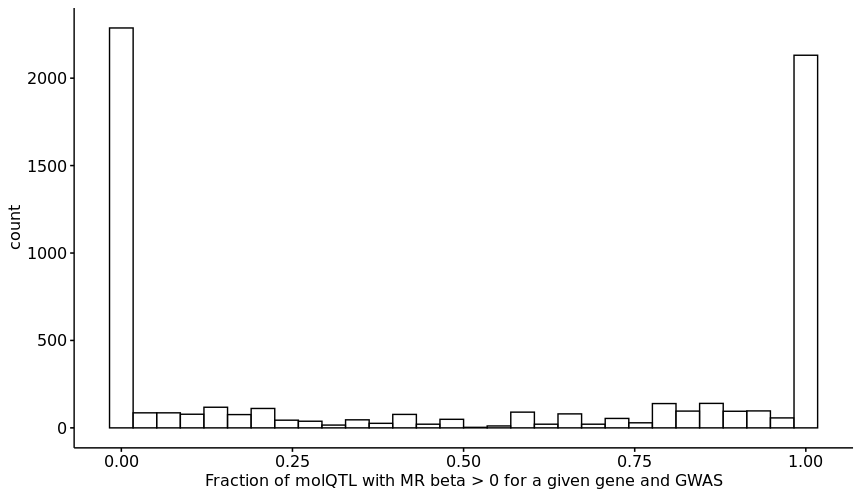

**Figure S5. Predicted direction of effect of gene expression on disease risk.** For a given gene and disease GWAS, we used mendelian randomization with molQTL (expression or protein QTL) as exposure to infer the impact of expression on disease risk (q-value < 0.05). We only included molQTL which colocalized with the local GWAS signal with a H4 posterior probability > 80%. For each gene and disease, we then calculated to fraction of colocalizing molQTL that were predicted to increase disease risk i.e. a fraction of 0 means that increased expression of gene X is predicted to decrease risk of disease Y across all molQTL datasets assessed. Conversely, a fraction of 1 means that increased expression of gene X is predicted to increase risk of disease Y across all molQTL assessed. We only include gene-GWAS pairs for which there were at least 5 colocalizing molQTL.

molQTL: Molecular quantitative trait loci; MR : Mendelian randomization; GWAS : Genome-wide association study

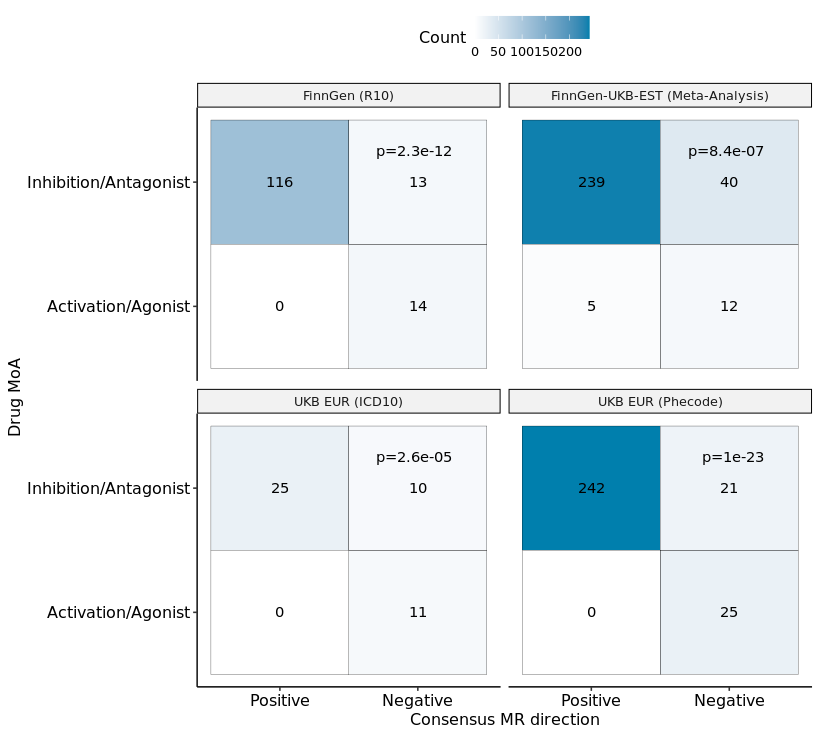

**Figure S6. Concordance between the predicted effect of gene expression on disease risk by mendelian randomization (MR) and mechanism of action (MoA) of approved drugs.** We retrieved information about targets, clinical trials, and drug MoA from the Citeline Pharmacogenomics dataset. This dataset was connected to GWAS phenotypes using EFO codes and a semantic similarity score > 0.7. The direction of effect of gene expression on disease risk was assessed by MR using molQTL as exposure (q-value < 0.05). Only molQTL colocalizing with local GWAS signal (H4 posterior probability > 80%) were included. A consensus direction was inferred if the MR direction of effect was consistent across > 75% of molQTL for a given gene and disease GWAS. A negative consensus MR direction suggests that increased gene expression leads to decreased disease risk. Therefore, an activator or agonist drug targeting this gene would be beneficial. Conversely, a positive consensus MR direction suggests that increased gene expression increases disease risk, and an inhibitor or antagonist drug would be beneficial. Reported P-values were calculated by Fisher exact test.

MR: Mendelian randomization; MoA: Mechanism of action

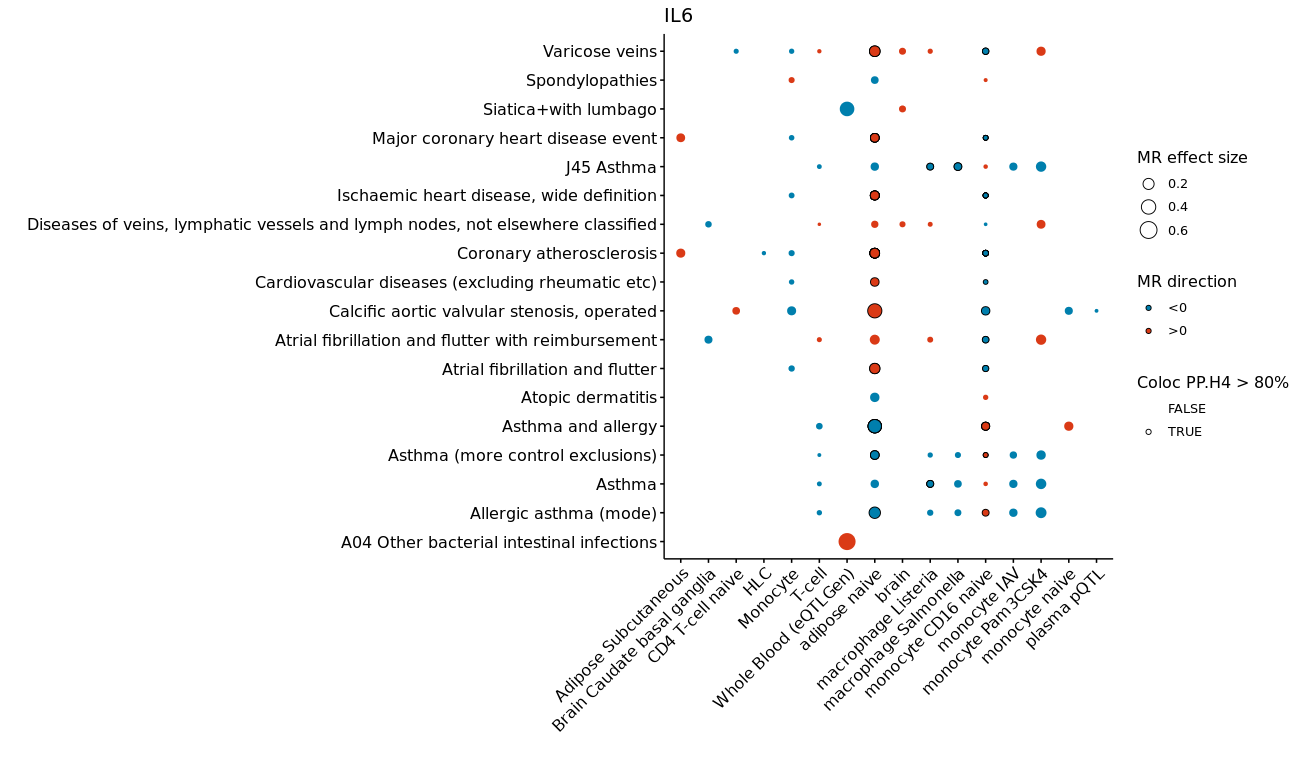

**Figure S7. Association between *IL6* and diseases, supported by MR, colocalization and ABC**. The figure shows tissues and cell-types with significant MR (q-value < 0.05) using *IL6* molQTL as exposure and diseases as outcome (red: positive effect size estimate [MR beta]; blue: negative effect size estimate). The size of the dots represents absolute MR effect size. Disease-molQTL pairs with a colocalization posterior probability > 80% are highlighted with a dark border.

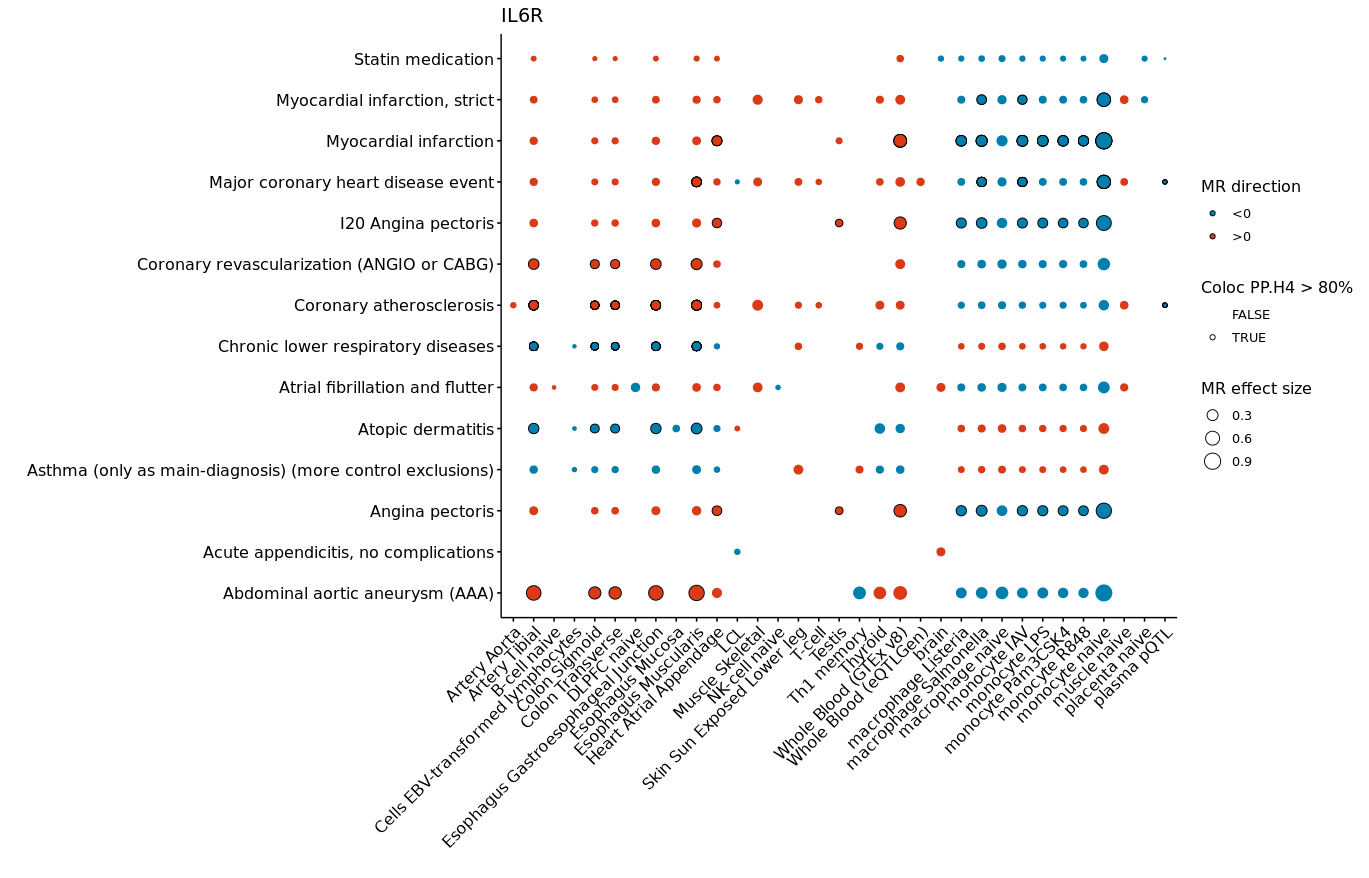

**Figure S8. Association between *IL6R* and diseases, supported by MR, colocalization and ABC**. The figure shows tissues and cell-types with significant MR (q-value < 0.05) using *IL6* molQTL as exposure and diseases as outcome (red: positive effect size estimate [MR beta]; blue: negative effect size estimate). The size of the dots represents absolute MR effect size. Disease-molQTL pairs with a colocalization posterior probability > 80% are highlighted with a dark border.
