## Additional File 4 for "Leveraging large-scale multi-omics to identify therapeutic targets from genome-wide association studies"

**Funding statements for molecular QTL data used in this manuscript.**

Genotype-Tissue Expression (GTEx) Project[1]

"This work was supported by the Common Fund of the Office of the Director, U.S. National Institutes of Health, and by NCI, NHGRI, NHLBI, NIDA, NIMH, NIA, NIAID, and NINDS through NIH contracts HHSN261200800001E (Leidos Prime contract with NCI: A.M.S., D.E.T., N.V.R., J.A.M., L.S., M.E.B., L.Q., T.K., D.B., K.R., and A.U.), 10XS170 (NDRI: W.F.L., J.A.T., G.K., A.M., S.S., R.H., G.Wa., M.J., M.Wa., L.E.B., C.J., J.W., B.R., M.Hu., K.M., L.A.S., H.M.G., M.Mo., and L.K.B.), 10XS171 (Roswell Park Cancer Institute: B.A.F., M.T.M., E.K., B.M.G., K.D.R., and J.B.), 10X172 (Science Care Inc.), 12ST1039 (IDOX), 10ST1035 (Van Andel Institute: S.D.J., D.C.R., and D.R.V.), HHSN268201000029C (Broad Institute: F.A., G.G., K.G.A., A.V.S., X.Li., E.T., S.G., A.G., S.A., K.H.H., D.T.N., K.H., S.R.M., and J.L.N.), 5U41HG009494 (F.A., G.G., and K.G.A.), and through NIH grants R01 DA006227-17 (University of Miami Brain Bank: D.C.M. and D.A.D.), Supplement to University of Miami grant DA006227 (D.C.M. and D.A.D.), R01 MH090941 (University of Geneva), R01 MH090951 and R01 MH090937 (University of Chicago), R01 MH090936 (University of North Carolina–Chapel Hill), R01MH101814 (M.M.-A., V.W., S.B.M., R.G., E.T.D., D.G.-M., and A.V.), U01HG007593 (S.B.M.), R01MH101822 (C.D.B.), U01HG007598 (M.O. and B.E.S.), U01MH104393 (A.P.F.), extension H002371 to 5U41HG002371 (W.J.K.), as well as other funding sources: R01MH106842 (T.L., P.M., E.F., and P.J.H.), R01HL142028 (T.L., Si.Ka., and P.J.H.), R01GM122924 (T.L. and S.E.C.), R01MH107666 (H.K.I.), P30DK020595 (H.K.I.), UM1HG008901 (T.L.), R01GM124486 (T.L.), R01HG010067 (Y.Pa.), R01HG002585 (G.Wa. and M.St.), Gordon and Betty Moore Foundation GBMF 4559 (G.Wa. and M.St.), 1K99HG009916-01 (S.E.C.), R01HG006855 (Se.Ka. and R.E.H.), BIO2015-70777-P, Ministerio de Economia y Competitividad and FEDER funds (M.M.-A., V.W., R.G., and D.G.-M.), la Caixa Foundation ID 100010434 under agreement LCF/BQ/SO15/52260001 (D.G.-M.), NIH CTSA grant UL1TR002550-01 (P.M.), Marie-Skłodowska Curie fellowship H2020 Grant 706636 (S.K.-H.), R35HG010718 (E.R.G.), FPU15/03635, Ministerio de Educación, Cultura y Deporte (M.M.-A.),R01MH109905, 1R01HG010480 (A.Ba.), Searle Scholar Program (A.Ba.), R01HG008150 (S.B.M.), 5T32HG000044-22, NHGRI Institutional Training Grant in Genome Science (N.R.G.), EU IMI program (UE7-DIRECT-115317-1) (E.T.D. and A.V.), FNS funded project RNA1 (31003A_149984) (E.T.D. and A.V.), DK110919 (F.H.), F32HG009987 (F.H.), Massachusetts Lions Eye Research Fund Grant (A.R.H.), Wellcome grant WT108749/Z/15/Z (P.F.), and European Molecular Biology Laboratory (P.F. and D.Z.).”

BLUEPRINT[2]

This study makes use of data generated by the Blueprint Consortium. A full list of the investigators who contributed to the generation of the data is available from www.blueprint-epigenome.eu. Funding for the project was provided by the European Union’s Seventh Framework Programme (FP7/2007- 2013) under grant agreement no 282510 – BLUEPRINT and the Canadian Institutes of Health Research (CIHR EP1-120608).

FUSION[3]

“This research was supported in part by National Institutes of Health Grants 1-ZIA-HG000024 (to F.S.C.), U01DK062370 (to M.B. and L.J.S.), and R00DK099240 (to S.C.J.P.); American Diabetes Association Pathway to Stop Diabetes Grant 1-14-INI-07 (to S.C.J.P.); Academy of Finland Grants 271961, 272741 (to M.L.), 258753 (to H.A.K.); European Molecular Biology Laboratory (to E.B.); UK Medical Research Council Grant MC_UU_00011/1 (to G.D.S.); and Wellcome Trust/Royal Society Grant 208806/Z/17/Z (to G.H.).”

GENCORD[4]

“Emmanouil T Dermitzakis was supported by grants from the European Research Council (260927), Swiss National Science Foundation (31003A_130342, CRSI33_130326) Louis-Jeantet Foundation, and the Blueprint Consortium. Stylianos E Antonarakis was supported by grants from the European Research Council (249968), Swiss National Science Foundation (144082), and the Blueprint Consortium.”

eQTLGen consortium [5]

This study makes use of data generated by the eQTLGen consortium (https://www.eqtlgen.org/ ): “This work is supported by a grant from the European Research Council (ERC, ERC Starting Grant agreement number 637640 ImmRisk), a VIDI grant (917.14.374) and a VICI grant from the Netherlands Organisation for Scientific Research (NWO) to L.F. This work has been supported by the European Regional Development Fund and the program Mobilitas Pluss (MOBTP108) to U.Võsa. The project was supported by the ‘De Drie Lichten’ foundation in the Netherlands with a grant to A.C. M.G.N. is supported by ZonMw grants 849200011 and 531003014 from the Netherlands Organisation for Health Research and Development, a VENI grant from the NWO (VI.Veni.191G.030) and a Jacobs Foundation research fellowship. H.Y. is funded by a Diabetes UK RD Lawrence fellowship (17/0005594). This project received funding from the ERC under the European Union’s Horizon 2020 research and innovation program (grant agreement no. 772376 (EScORIAL)) to J.H.V. T.E. and A.K. were supported by the Estonian Research Council grant PRG (PRG1291). A.Battle was supported by NIH grant R01MH109905, NIH grant R01HG008150 (NHGRI; Non-Coding Variants Program) and NIH grant R01MH101814 (NIH Common Fund; GTEx Program). M.G.P.v.d.W. was funded by the Nederlandse Organisatie voor Wetenschappelijk onderzoek, NWO-Veni 192.029. This work was supported by NIH grants R21ES024834 (B.Pierce), R01ES020506 (B.Pierce), R01ES023834 (B.Pierce), R35ES028379 (B.Pierce) and R01CA107431 (H.A.). This work was supported by the Sigrid Juselius Foundation (J.Kettunen) and funds from the Academy of Finland (grant numbers 297338 and 307247) (J.Kettunen) and the Novo Nordisk Foundation (grant number NNF17OC0026062) (J.Kettunen). S.Ripatti was supported by the Academy of Finland Centre of Excellence in Complex Disease Genetics (grant no. 312062). M.G. was supported by EU Horizon 2020 (grant 733100 for SYSCID) and a grant from the Excellence of Science (FNRS and FWO) (grant no. 30770923). We acknowledge support from the BBMRI-NL (Biobanking and Biomolecular Resources Research Infrastructure 184.021.007 and 184.033.111), Spinozapremie (NWO 56-464-14192), the ERC (ERC Advanced 230374) and the KNAW Academy Professor Award (PAH/6635) to D.I.B. G.H. works in a unit that receives funding from the UK MRC (MC_UU_12013/1&2&5) and the University of Bristol. S.B. was supported by the Swiss National Science Foundation (310030-152724). B.M.P. was supported by CHARGE infrastructure grant number HJ105756 for the HVH cohort. This work was supported by the German Federal Ministry of Education and Research (BMBF) within the framework of the e:Med research and funding concept (grant 01ZX1906B) and by LIFE (Leipzig Research Center for Civilization Diseases), Universität Leipzig (which is funded by the European Union, by the European Regional Development Fund and by the Free State of Saxony within the framework of the excellence initiative to H.K. and M.Scholz). We thank the UMCG Genomics Coordination Center, the MOLGENIS team, the UG Center for Information Technology and the UMCG research IT program and their sponsors, in particular the BBMRI-NL for data storage, high-performance computing and web hosting infrastructure. The BBMRI-NL is a research infrastructure financed by the NWO (grant number 184.033.111).”

Schmiedel 2018 (DICE)

“This work was funded by the William K. Bowes Jr Foundation (P.V.) and NIH grants R24AI108564 (P.V., B.P., A.R., M.K.), S10RR027366 (BD FACSAria II), and S10OD016262 (Illumina HiSeq 2500).”

CAP[6]

“This project was supported by NIH U19 HL069757, AHA 15POST21880006, NIH P50 GM115318, and NIH R01 HL139902. This study was also supported in part by the NIH Pharmacogenomics Research Network (PGRN) RNA Sequencing Project, the National Center for Advancing Translational Sciences, CTSI grant ULTR001881, and the National Institute of Diabetes and Digestive and Kidney Disease Diabetes Research Center (DRC) grant DK063491 to the Southern California Diabetes Endocrinology Research Center. The funders had no role in study design, data collection and analysis, data interpretation, or manuscript writing.”

Alasoo 2018[7]

“This work was supported by Wellcome Trust grant WT098051 (G.D. and D.J.G.). K.A. was supported by a PhD fellowship from the Wellcome Trust (WT099754/Z/12/Z) and a postdoctoral fellowship from the Estonian Research Council (MOBJD67). The iPSC lines were generated at the Wellcome Trust Sanger Institute, under the Human Induced Pluripotent Stem Cell Initiative funded by a strategic award (WT098503) from the Wellcome Trust and Medical Research Council.”

HipSci [8, 9]

This study makes use of data generated by the HipSci Consortium, funded by The Wellcome Trust and the MRC: “This work was funded with a strategic award from the Wellcome Trust and UK Medical Research Council (WT098503). Work at the Wellcome Trust Sanger Institute was further supported by Wellcome Trust grant WT090851. H.K. is supported by a MRC eMedLab Medical Bioinformatics career development award from the UK Medical Research Council (MR/L016311/1). F.M.W. acknowledges financial support from the Department of Health via the NIHR Biomedical Research Centre award to Guy’s & St Thomas’ National Health Service Foundation Trust in partnership with King’s College London and King’s College Hospital NHS Foundation Trust. “

Peng 2018[10]

“KH is partially supported by National Natural Science Foundation of China (Grant No. 21477087, 91643201, 21876134), and by Ministry of Science and Technology of China (Grant No. 2016YFC0206507). SP, MAD, CM, JC, KH are partially supported by National Institute of Environmental Health Sciences (NIH-NIEHS R01ES022223, R01ES022223-03S1, 1R01ES029212-01).”

Quach 2016[11]

“KH is partially supported by National Natural Science Foundation of China (Grant No. 21477087, 91643201, 21876134), and by Ministry of Science and Technology of China (Grant No. 2016YFC0206507). SP, MAD, CM, JC, KH are partially supported by National Institute of Environmental Health Sciences (NIH-NIEHS R01ES022223, R01ES022223-03S1, 1R01ES029212-01).”

TwinsUK[12]

“TwinsUK is funded by the Wellcome Trust, Medical Research Council, European Union, the National Institute for Health Research (NIHR)-funded BioResource, Clinical Research Facility and Biomedical Research Centre based at Guy’s and St Thomas’ NHS Foundation Trust in partnership with King’s College London.”

Nedelec 2016[13]

“This study was funded by grants to L.B.B. from the Canadian Institutes of Health Research (301538 and 232519), the Human Frontiers Science Program (CDA-00025/2012), and the Canada Research Chairs Program (950-228993). We thank Calcul Québec and Compute Canada for providing access to the supercomputer Briaree from the University of Montreal. Y.N. was supported by a fellowship from the Réseau de Médecine Génétique Appliquée (RMGA); J.S. by a fellowship from the Fonds de recherche du Québec-Nature et technologies (FRQNT), and G.B. and J.S. by a fellowship from the Fonds de recherche du Québec-Santé (FRQS).”

Geuvadis[14]

This study makes use of data generated by the Geuvadis Consortium (<https://www.internationalgenome.org/data-portal/data-collection/geuvadis>): “This project was funded by the European Commission 7th Framework Program (FP7) (261123; GEUVADIS); the Swiss National Science Foundation (130326, 130342), the Louis Jeantet Foundation, and ERC (260927) (E.T.D.); NIH-NIMH (MH090941) (E.T.D., M.I.M., R.G.); Spanish Plan Nacional SAF2008-00357 (NOVADIS), the Generalitat de Catalunya AGAUR 2009 SGR-1502, and the Instituto de Salud Carlos III (FIS/FEDER PI11/00733) (X.E.); Spanish Plan Nacional (BIO2011-26205) and ERC (294653) (R.G.); ESGI, READNA (FP7 Health-F4-2008-201418), Spanish Ministry of Economy and Competitiveness (MINECO) and the Generalitat de Catalunya (I.G.G.); DFG Cluster of Excellence Inflammation at Interfaces, the INTERREG4A project HIT-ID, and the BMBF IHEC project DEEP SP 2.3 (P.Ro.); German Centre for Cardiovascular Research (DZHK) and the German Ministry of Education and Research (01GR0802, 01GM0867, 01GR0804, 16EX1020C) (T.M.); EurocanPlatform (FP7 260791), ENGAGE and CAGEKID (241669) (A.B.); FP7/2007-2013, ENGAGE project, HEALTH-F4-2007-201413, and the Centre for Medical Systems Biology within the framework of The Netherlands Genomics Initiative (NGI)/Netherlands Organisation for Scientific and Research (NWO) (P.AC.H and G.-J.v.O.); The Swedish Research Council (C0524801, A028001) and the Knut and Alice Wallenberg Foundation (2011.0073) (A.-C.S.); The Swiss National Science Foundation (127375, 144082) and ERC (249968) (S.E.A.); Instituto de Salud Carlos III (FIS/FEDER PS09/02368) (A.C.); German Federal Ministry of Education and Research (01GS08201) (R.S.); Max Planck Society (H.L.); Wellcome Trust (WT085532) and the European Molecular Biology Laboratory (P.F.); ENGAGE, Wellcome Trust (081917, 090367, 090532, 098381), and Medical Research Council UK (G0601261) (M.I.M.); Wellcome Trust Centre for Human Genetics (090532/Z/09/Z, 075491/Z/04/B), Wellcome Trust (098381, 090367, 076113, 083270), the WTCCC2 project (085475/B/08/Z, 085475/Z/08/Z), Royal Society Wolfson Merit Award, Wellcome Trust Senior Investigator Award (095552/Z/11/Z) (P.D.); EMBO long-term fellowship EMBO-ALTF 2010-337 (H.K.); NIH-NIGMS (R01 GM104371) (D.G.M.); Marie Curie FP7 fellowship (O.S.); Scholarship by the Clarendon Fund of the University of Oxford, and the Nuffield Department of Medicine (M.A.R.); EMBO long-term fellowship ALTF 225-2011 (M.R.F.); Emil Aaltonen Foundation and Academy of Finland fellowships (T.L.).”

ROSMAP[15]

“Study data were provided by the Rush Alzheimer s Disease Center, Rush University Medical Center, Chicago. Data collection was supported through funding by NIA grants P304G10161, R014G15819, R014G17917, R01AG30!46, R014G36836, U014G32984, U014G46152, the Illinois Deparhnenr of Public Health, and the Translational Genomics Research Institute.”

CommonMind[16]

“Data were generated as part of the CommonMind Consortium supported by funding from Takeda Pharmaceuticals Company Limited, F. Hoffmann-La Roche Ltd and NIH grants U01MH116442, R01MH085542, R01MH093725, P50MH066392, P50MH080405, R01MH097276, RO1-MH-075916, P50M096891, P50MH084053S1, R37MH057881, AG02219, AG05138, MH06692, R01MH110921, R01MH109677, R01MH109897, U01MH103392, and contract HHSN271201300031C through IRP NIMH. Brain tissue for the study was obtained from the following brain bank collections: the Mount Sinai NIH Brain and Tissue Repository, the University of Pennsylvania Alzheimer’s Disease Core Center, the University of Pittsburgh NeuroBioBank and Brain and Tissue Repositories, and the NIMH Human Brain Collection Core. CMC Leadership: Panos Roussos, Joseph Buxbaum, Andrew Chess, Schahram Akbarian, Vahram Haroutunian (Icahn School of Medicine at Mount Sinai), Bernie Devlin, David Lewis (University of Pittsburgh), Raquel Gur, Chang-Gyu Hahn (University of Pennsylvania), Enrico Domenici (University of Trento), Mette A. Peters, Solveig Sieberts (Sage Bionetworks), Thomas Lehner, Stefano Marenco, Barbara K. Lipska (NIMH). Rhesus Macaque tissue was provided by Scott Hemby through the Stanley Medical Research Institute for Funding for Non-Human Primate Research; and funded by NIMH grant R01MH074313. J.B. was supported in part by NARSAD Young Investigator Grant 27209 from the Brain & Behavior Research Foundation. G.E.H. was supported in part by NARSAD Young Investigator Grant 26313 from the Brain & Behavior Research Foundation. This work was supported in part through the computational resources and staff expertise provided by the Department of Scientific Computing at the Icahn School of Medicine at Mount Sinai.”

Steinberg 2021[17]

“This work was funded by the Wellcome Trust (206194). M.J.C. was funded through a Medical Research Council Centre for Integrated Research into Musculoskeletal Ageing grant (148985). R.A.B. and the Human Research Tissue Bank are supported by the NIHR Cambridge Biomedical Research Centre. J.H.D.B. and G.R.W. are funded by a Wellcome Trust Strategic Award (101123), a Wellcome Trust Joint Investigator Award (110140 and 110141) and a European Commission Horizon 2020 Grant (666869, THYRAGE). A.W.M. receives funding from Versus Arthritis; Tissue Engineering and Regenerative Therapies Centre (21156).”

iPSCORE[18]

" This work was supported in part by a California Institute for Regenerative Medicine (CIRM) grant GC1R-06673 (to K.A.F.), TR3-05687 (to E.A.), and NIH grants HG008118 (to K.A.F.), HL107442 (to K.A.F., S.M.E., G.W.Y., N.C.C., and J.C.I.B.), DK105541 (to K.A.F.) and DK112155 (to K.A.F.); The following individuals were partially supported by NIH Supplements: A.D.A. (HL107442), M.G. (HG008118), and V.B. (EY021237). C.Z. was supported by CIRM Bridges TBI-01186 grant and K.J.F. and M.C. by CIRM Bridges II EDUC2-08375 grant awarded to California State University San Marcos (to Bianca R. Mothé). The Leona M. and Harry B. Helmsley Charitable Trust (2012-PG-MED002) (J.C.I.B.), Universidad Catolica San Antonio de Murcia (UCAM) and the G. Harold and Leila Y. Mathers Charitable Foundation (J.C.I.B.), the NIH grant UL1TR000100 of CTSA funding prior to August 13, 2015 and grant UL1TR001442 of CTSA funding beginning August 13, 2015 and beyond; J.D.G. and C.D. are supported in part by an institutional award to the UCSD Genetics Training Program from the NIGMS (T32GM008666); C.D. is supported in part by CIRM Interdisciplinary Stem Cell Training Program at UCSD II (TG2-01154); D.A.J. and M.K.R.D. are supported in part by NIH National Library of Medicine Training Grant (4T15LM011271). W.W.G. is supported in part by U.S. Department of Health and Human Services training grant (5T32GM008806-15). P.B. is supported in part by the Swiss National Science Foundation (P2LAP3-155105 and P300PA-167612); HumanCoreExome array data were generated at the UCSD IGM Genomics Center with support from NIH grant P30CA023100. R.M.W. and J.F.L. were supported by CIRM Award for Tools and Technologies (RT3-07655). F.-J.M. and B.M.S. were supported by grants from the BMBF (13GW0128A and 01GM1513D) and from the Deutsche Forschungsgemeinschaft (German Research Foundation DFG MU 3231/3-1).”

Braineac2[19]

"Mina Ryten, David Zhang, and Karishma D’Sa were supported by the UK Medical Research Council (MRC) through the award of Tenure-track Clinician Scientist Fellowship to Mina Ryten (MR/N008324/1). Sebastian Guelfi was supported by Alzheimer’s Research UK through the award of a PhD Fellowship (ARUK-PhD2014-16). Regina Reynolds was supported through the award of a Leonard Wolfson Doctoral Training Fellowship in Neurodegeneration.”

BrainSeq[20]

"This work was supported by the funding from Lieber Institute for Brain Development and the Maltz Research Laboratories and partially supported by NIH R21MH109956 (A.E.J.) and Consejo Nacional de Ciencia y Tecnología México 351535 (L.C.-T.). The Genotype-Tissue Expression (GTEx) Project was supported by the Common Fund of the Office of the Director of the National Institutes of Health. Additional funds were provided by the NCI, NHGRI, NHLBI, NIDA, NIMH and NINDS. Donors were enrolled at Biospecimen Source Sites funded by NCI/SAIC-Frederick, Inc. (SAIC-F) subcontracts to the National Disease Research Interchange (10XS170), Roswell Park Cancer Institute (10XS171) and Science Care, Inc. (X10S172). The Laboratory, Data Analysis, and Coordinating Center (LDACC) was funded through a contract (HHSN268201000029C) to The Broad Institute, Inc. Biorepository operations were funded through an SAIC-F subcontract to the Van Andel Institute (10ST1035). Additional data repository and project management were provided by SAIC-F (HHSN261200800001E). The Brain Bank was supported by supplements to University of Miami grants DA006227 and DA033684 and to contract N01MH000028. Statistical methods development grants were made to the University of Geneva (MH090941 and MH101814), the University of Chicago (MH090951, MH090937, MH101820, MH101825), the University of North Carolina - Chapel Hill (MH090936 and MH101819), Harvard University (MH090948), Stanford University (MH101782), Washington University St Louis (MH101810) and the University of Pennsylvania (MH101822). The data used for the analyses described in this manuscript were obtained from dbGaP accession number phs000424.v6.p1 on October 6, 2015. Data were generated as part of the CommonMind Consortium supported by funding from Takeda Pharmaceuticals Company Limited, F. Hoffman-La Roche Ltd and NIH grants R01MH085542, R01MH093725, P50MH066392, P50MH080405, R01MH097276, R01-MH-075916, P50M096891, P50MH084053S1, R37MH057881, R37MH057881S1, HHSN271201300031C, AG02219, AG05138 and MH06692. Brain tissue for the study was obtained from the following brain bank collections: the Mount Sinai NIH Brain and Tissue Repository, the University of Pennsylvania Alzheimer’s Disease Core Center, the University of Pittsburgh NeuroBioBank and Brain and Tissue Repositories and the NIMH Human Brain Collection Core.”

PhLiPS[21]

“This work was supported in part by the Howard Hughes Medical Institute Medical Research Fellows Program (A.R.), grant R01-GM104464 from the NIH (K.M. and T.S.M.), grants R01-HL118744 and R01-DK099571 from the NIH (K.M.), the Harvard Stem Cell Institute (K.M.), grant U01-HG006398 from the NIH (D.J.R., E.E.M., and S.A.D.), RC2-HL101864 from the NIH (D.J.R.), grant UL1-TR000003 from the NIH/NCATS (D.J.R.), and R01-MH101822 from the NIH (C.D.B.).”

Young 2019[22]

"R.F. was supported by funding from the UK Multiple Sclerosis Society (MS50), the Adelson Medical Research Foundation and a core support grant from the Wellcome Trust and MRC to the Wellcome Trust-Medical Research Council Cambridge Stem Cell Institute (203151/Z/16/Z). A.Y. is supported by a Wellcome Trust Clinicians PhD Fellowship (RRZD/029). All data for this study were generated under Open targets project OTAR039. N.K. and D.J.G. were funded by the Wellcome Trust grant WT206194.”

Schwartzentruber 2018[23]

"The iPSC lines were generated under the Human Induced Pluripotent Stem Cell Initiative (HIPSCI) funded by a grant from the Wellcome Trust and Medical Research Council (WT098503), supported by the Wellcome Trust (WT098051) and the NIHR/Wellcome Trust Clinical Research Facility. HIPSCI funding was used for sensory neuron RNA-sequencing. We acknowledge Life Science Technologies Corporation as the provider of Cytotune. Pfizer Neuroscience (Pfizer Ltd.) funded neuronal differentiation, functional assays, single-cell RNA-sequencing, and collection and sequencing of dorsal root ganglion samples.”

Van de Bunt 2015[24]

“MvdB is supported by a Novo Nordisk postdoctoral fellowship run in partnership with the University of Oxford. ALG is a Wellcome Trust Senior Research Fellow in Basic Biomedical Science (095010/Z/10/Z). MIM is a Wellcome Trust Senior Investigator (WT098381) and a National Institute of Health Research Senior Investigator. PEM holds the Canada Research Chair in Islet Biology. This work was supported in part in Oxford, UK, by grants from the Medical Research Council (MRC; MR/L020149/1) and National Institutes of Health (NIH; R01 MH090941), and in Edmonton, Canada, by operating grants to PEM from the Canadian Institutes of Health Research (CIHR; MOP244739) and the ADI/Johnson & Johnson Diabetes Research Fund. Human islet isolations at the Alberta Diabetes Institute IsletCore were funded by the Alberta Diabetes Foundation and the University of Alberta. The National Institute for Health Research, Oxford Biomedical Research Centre funded islet provision at the Oxford Human Islet Isolation facility. The funders had no role in study design, data collection and analysis, decision to publish, or preparation of the manuscript.”

Sun 2018[25]

“We thank INTERVAL study participants; staff at recruiting NHSBT blood donation centres; and the INTERVAL Study Co-ordination team, Operations Team (led by R. Houghton and C. Moore) and Data Management Team (led by M. Walker). This research was supported as follows. The Cardiovascular Epidemiology Unit at the University of Cambridge: UK MRC (G0800270), BHF (SP/09/002), UK NIHR Cambridge Biomedical Research Centre, ERC (268834), and European Commission Framework Programme 7 (HEALTH-F2-2012-279233); B.B.S.: Cambridge School of Clinical Medicine MB-PhD programme and MRC/Sackler Prize PhD Studentship (MR/K50127X/1); J.E.P.: a BHF Cambridge Centre of Excellence Research Fellowship [RE/13/6/30180] and a UK Research Innovation Fellowship (MR/S004068/1). P.S.: a Rutherford Fund Fellowship (MR/S003746/1); D.S.P. and D.S.: the Wellcome Trust (105602/Z/14/Z); N.S.: the Wellcome Trust (WT098051 and WT091310), the EU FP7 (EPIGENESYS 257082 and BLUEPRINT HEALTH-F5-2011-282510); J.A.T: the Wellcome Trust (091157) and JDRF (9-2011-253); K.S.: the Biomedical Research Program at Weill Cornell Medicine in Qatar via the Qatar Foundation; J.D.: BHF Professor, European Research Council Senior Investigator, and NIHR Senior Investigator; the INTERVAL study: NHSBT (11-01-GEN) and the NIHR-BTRU in Donor Health and Genomics (NIHR BTRU-2014-10024) at the University of Cambridge in partnership with NHSBT. Data analysis was partly supported by the Cambridge Substantive Site of Health Data Research UK. This study was partially funded by Merck Sharp & Dohme Corp.”
